# A Metapopulation Network Model for the Spreading of SARS-CoV-2: Case Study for Ireland^⋆^

**DOI:** 10.1101/2020.06.26.20140590

**Authors:** Rory Humphries, Mary Spillane, Kieran Mulchrone, Sebastian Wieczorek, Micheal O’Riordain, Philipp Hövel

## Abstract

We present preliminary results on an all-Ireland network modelling approach to simulate the spreading the Severe Acute Respiratory Syndrome CoronaVirus 2 (SARS-CoV-2), commonly known as the *coronavirus*. In the model, nodes correspond to locations or communities that are connected by links indicating travel and commuting between different locations. While this proposed modelling framework can be applied on all levels of spatial granularity and different countries, we consider Ireland as a case study. The network comprises 3440 *electoral divisions* (EDs) of the Republic of Ireland and 890 *superoutput areas* (SOAs) for Northern Ireland, which corresponds to local administrative units below the NUTS 3 regions. The local dynamics within each node follows a phenomenological SIRX compartmental model including classes of Susceptibles, Infected, Recovered and Quarantined (X) inspired from Science **368**, 742 (2020). For better comparison to empirical data, we extended that model by a class of Deaths. We consider various scenarios including the 5-phase roadmap for Ireland. In addition, as proof of concept, we investigate the effect of dynamic interventions that aim to keep the number of infected below a given threshold. This is achieved by dynamically adjusting containment measures on a national scale, which could also be implemented at a regional (county) or local (ED/SOA) level. We find that – in principle – dynamic interventions are capable to limit the impact of future waves of outbreaks, but on the downside, in the absence of a vaccine, such a strategy can last several years until herd immunity is reached.

---

On 30th January, 2020, the World Health Organisation (WHO) declared the newly emergent novel coronavirus pathogen, Severe Acute Respiratory Syndrome CoronaVirus 2 (SARS-CoV-2), to be a *public health emergency of international concern* [41, 42]. This characterisation was later updated to pandemic status on 11th March 2020 [43]. Following its initial detection in a cluster of patients experiencing acute respiratory symptoms in the city of Wuhan, in the Hubei province of China [13], the spread of SARS-CoV-2, which causes the coronavirus disease 2019 (COVID-19), to 218 countries and territories in the months since has led to unprecedented social and economic disruption [18, 45]. Despite widespread efforts to contain the virus through a series of non-pharmaceutical interventions, global figures show reported cases standing at over 63 million, with deaths from the virus totalling 1.5 million up to late November 2020 [17, 25]. It is clear that there is an urgent and ongoing need to understand how the virus spreads in order to effectively protect populations.

As of yet there are no significantly effective pharmaceutical measures available that prevent the spread of the disease. Vaccination programmes commenced on 14th December, 2020 and in the first month, approximately 35 million doses have been administered in 49 countries worldwide [7]. Immunisation of billions of people around the world will be a logistical tour de force, and the time frame for a significant level of coverage is still on the order of many months or more. Until such measures are widely implemented, the best available mitigation strategies include the use of face masks, sanitisation, and social distancing. Authorities have implemented lockdowns to promote these measures. Mathematical models are invaluable in providing us with an insight into the processes by which the disease spreads [1, 8, 29–31, 33] and these models can help decide on the best course of action to take in the form of non-pharmaceutical interventions [4, 34–36].

The model proposed in this discussion paper and the numerical results arising from it are to be understood as a contribution to the scientific and public discussion of possible spreading scenarios and the impact of intervention measures including realistic levels of compliance. We will clearly state the model ingredients and assumptions and elaborate on conclusions that can be drawn based on these limitations; some of which are unlikely for any practical purpose, but worth exploring, e.g., for worst-case scenarios.

We start with an introduction of the model equations for the local and networked dynamics in sections 1 and 2, respectively. This includes a discussion of the local dynamics from a geometric point of view, where we sketch the trajectories and equilibria in phase space. Then, we continue with a case study of Ireland in section 3 including realistic mobility data and compliance levels with respect to movement restrictions. Besides an unlikely, worst-case scenario, we consider different levels of increased awareness during the post-lockdown period in section 3.2. In addition, in section 3.3, we explore effects of dynamic interventions to keep the prevalence below a given level. These dynamic lockdowns are considered at different levels of spatial granularity.

## 1. Local Dynamics

We start with the model for the local, well mixed dynamics at a single node as the basis for the network model:

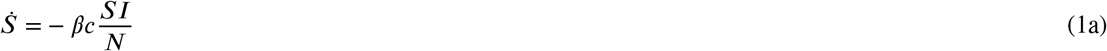

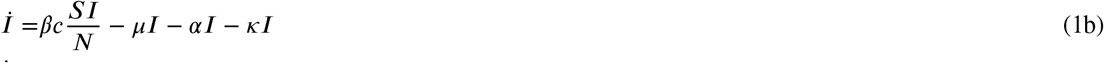

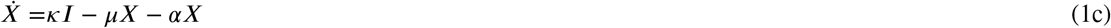

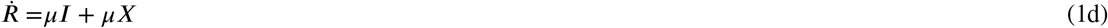

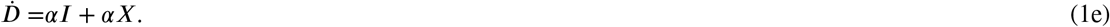

The above model is a simple extension of the original Kermack and McKendrick SIR model [26], inspired by Ref. [31]. There are five compartments, *N* individuals, and each individual may belong to just one compartment at any time:

**Susceptible** *S*(*t*) Individuals who have not been infected by the disease.

**Infected** *I*(*t*) Individuals who are infected with the disease and are capable of transmission.

**Quarantined** *X*(*t*) Individuals who are infected with the disease, but are quarantined or self-isolating and thus not transmitting.

**Recovered** *R*(*t*) Individuals who have recovered from the disease and are considered immune.

**Dead** *D*(*t*) Individuals who have died from the disease.

The above system of ordinary differential equations (ODEs) assume homogeneous mixing of the population, which be a good approximation for small communities, and small number of deaths. The parameters of the ODE model (1) refer to the rates of moving from one compartment to another and population size:

*β* probability of infection on contact.

*c* rate of contact/mixing.

*µ* rate of recovery.

*κ* rate of moving from infected to quarantined.

*α* rate of death.

*N* total population size *N* = *S*(*t*) + *I*(*t*) + *X*(*t*) + *R*(*t*) + *D*(*t*) = const..

Much insight into the network dynamics can be gained from geometric analysis of the local, single-node dynamics. In the (*S, I, X, R, D*) phase space, system (1) has a whole family of stationary solutions (equilibria). They are given by

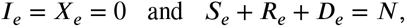

correspond to disease-free conditions, and form a triangular surface shown in Fig. 1. This family includes the important pre-disease equilibrium

**Figure 1:**
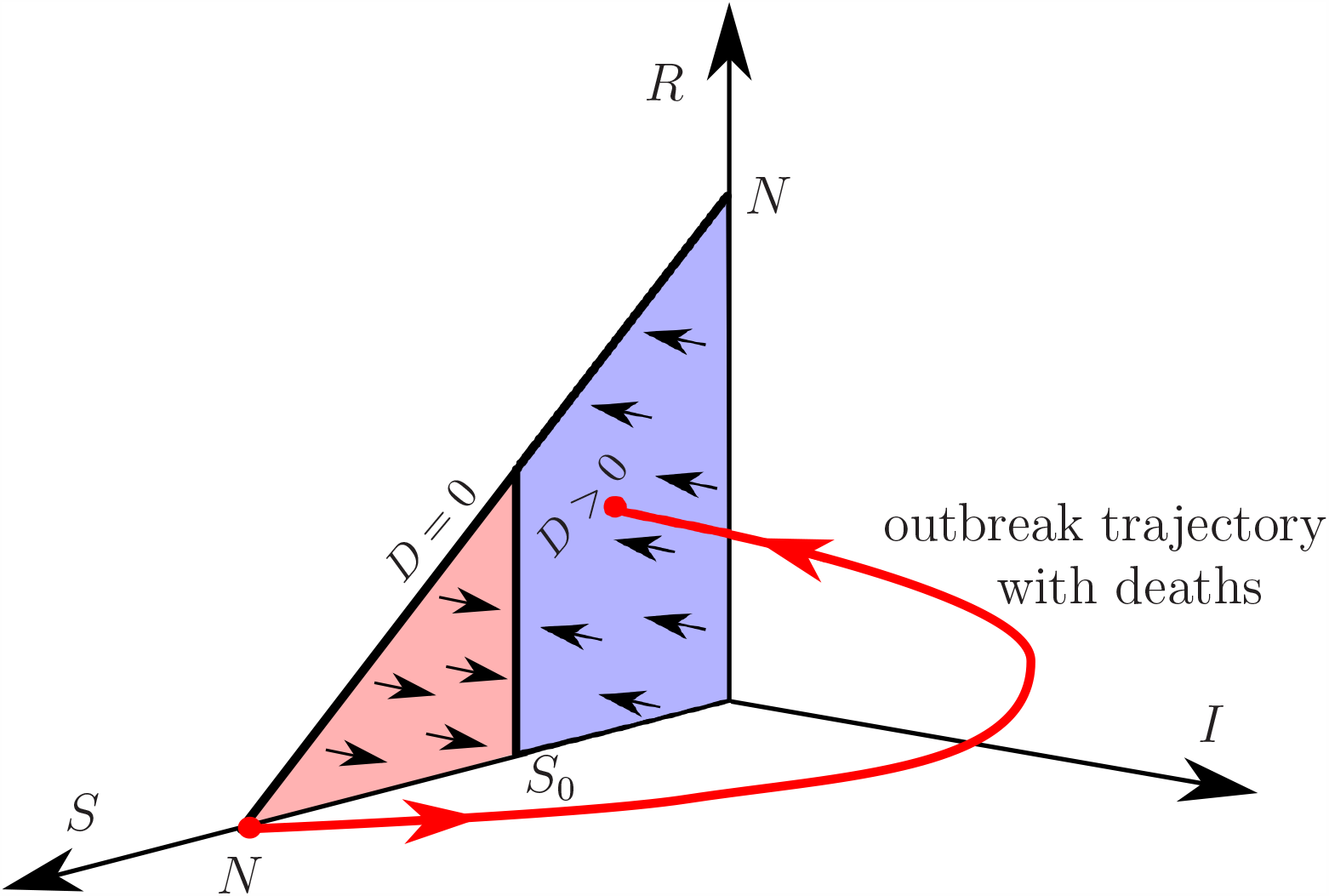
Sketch of the outbreak dynamics in the local SIXRD model (1) for *R*_0_ > 1, shown as a (red) trajectory from the disease-free equilibrium *S*_e_ = *N* to one of the stable equilibria with 0 < *S*_e_ < *S*_0_ in the projection of the (*S, I, X, R, D*) phase space onto the (*S, I, R*) subspace. *S*_0_ is the herd immunity threshold separating the triangular family of equilibria into the (pink) unstable and (blue) stable parts.

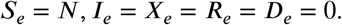

To discuss stability of equilibria, it is convenient to introduce the *basic reproduction number*:

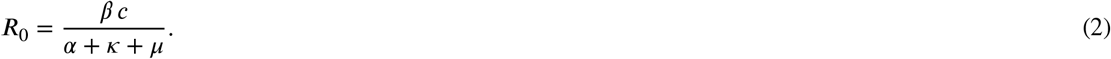

When 0 < *R*_0_ < 1, all equilibria are stable, meaning that no disease outbreaks can occur. When *R*_0_ > 1, the pre-disease equilibrium turns unstable, and disease outbreaks become possible. To be more specific, the family of equilibria is divided into the (pink) unstable part with *S*_0_ < *S*_e_ ≤ *N*, which contains the pre-disease equilibrium, and the (blue) stable part with 0 ≤ *S*_e_ < *S*_0_, shown in Fig. 1. The instability threshold separating the two parts is given by

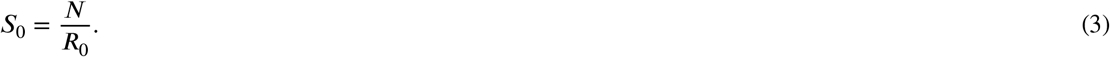

Guided by these observations, we define *herd immunity*, whether achieved naturally or by means of vaccination, in terms of stable equilibria as follows:

### Definition.

*In the local SIXRD model* (1), *we define* herd immunity *as a disease-free state of the system such that a small increase in infected I decays monotonically to zero* (*no outbreak*):

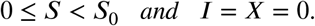

*In other words, herd immunity is represented by a stable equilibrium. We call S*_0_ = *N*/*R*_0_ *the herd immunity threshold*.

In Fig. 1, a disease outbreak is depicted as the (red) trajectory from the pre-disease equilibrium, giving a peak in infected *I*(*t*) around the herd immunity threshold *S*_0_, to one of the (blue) herd immunity equilibria that lie below *S*_0_. The lower the *S*_0_, the higher the peak in *I*(*t*). Thus, bringing the herd immunity threshold *S*_0_ closer to *N* for a period of time and then back to its original position, e.g., via imposing temporary public-health restrictions, can modify the outbreak trajectory so that ultimate herd immunity is achieved in a controlled way, that is with a peak in *I*(*t*) that remains below some desired level.

## 2. Network Model

While the ODE model (1) can be justified for small communities, it fails at capturing the dynamics that occurs at large spatial scales and cannot account for heterogeneities in a population and interaction between metapopulations via mixing/commuting [5]. For this reason, we develop a network framework by splitting the population up into a number of smaller communities based on their geographic location. Ideally, these communities are small enough to justify a well mixed ODE model at the local level. Every community corresponds to a node in the network. Two nodes are connected by a link, which represent the number of people travelling between the involved communities. The number of people travelling can vary with time or remain static. The resulting network model is given by

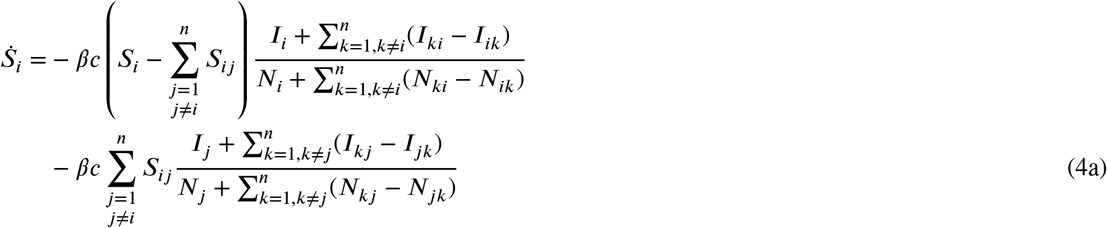

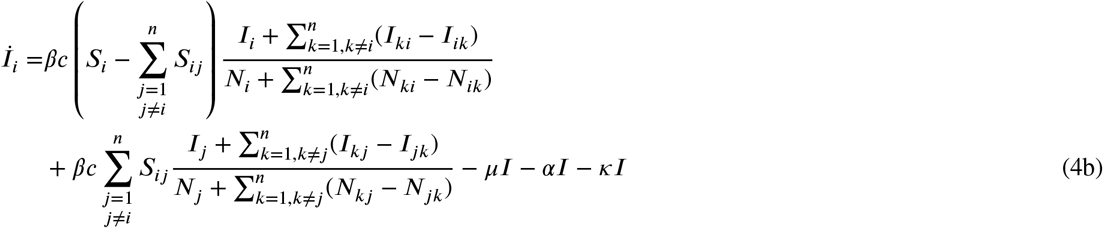

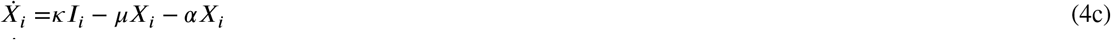

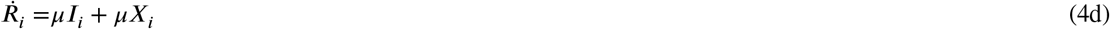

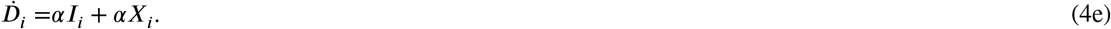

For notational convenience, the compartments are now labelled by a subscript, i.e., *S*_*i*_, *I*_*i*_, *X*_*i*_, *R*_*i*_, and *D*_*i*_, to denote number of individuals belonging to the respective compartments of *i*-th node in the netw k (*S*_*i*_ + *I*_*i*_ + *X*_*i*_ + *R*_*i*_ + *D*_*i*_ = *N*_*ij*_, *i* = 1, …, *n*). Similarly, the number of individuals travelling from node *i* to node belonging to a given compartment is denoted by *S*_*ij*_, *I*_*ij*_, *X*_*ij*_, *R*_*ij*_, and *D*_*ij*_ resulting in *N*_*ij*_ = *S*_*ij*_ + *ij*_*ij*_ + *X*_*ij*_ + *R*_*ij*_ + *D*_*ij*_. The proposed modelling framework accounts for travel as commuting pattern, that is, individuals return to their original location (cf. Appendix D). The local dynamics is included via the single-subscript quantities. Hence, *S*_*ii*_ = *I*_*ii*_ = *X*_*ii*_ = *R*_*ii*_ = *D*_*ii*_ = 0. Note that the network model includes interactions between nodes only in the susceptible and infected compartments. Accordingly, we account for the number of (i) individuals that remain in their home node and (ii) those who travel to other nodes in the first and second term of Eqs. (4a) and (4b), respectively. For example, the term 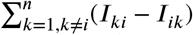 in Eq. (4a) refers to the net flux of infected individuals, that is, the difference between individuals leaving node *i* to *k* and arriving from node. *k* to *i*

Given a number of travellers *N*_*ij*_ between nodes, the network model (4) assumes that the number of travellers belonging to the respective compartments is determined by a relation such as 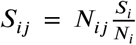. Thus, the number of travelling susceptibles is proportional to the number of susceptibles in the home node. The number of travelling individuals in the other compartments is computed in the same fashion. *N*_*ij*_ may be randomly generated at each time step. The distance travelled between locations often follows a power-law or heavy-tail distribution and can be described by a gravity or radiation population model [9, 10, 20, 37–39]. Alternatively, empirical movement data can inform the model. From these data one can either build a static network using, e.g., the jump-size distributions, or randomly generate a network at each time-step. Note that the proposed network model does not account for any importation from outside the considered network.

For the fully networked case [cf. Eqs. (4)], the basic reproduction number *R*_0_ can be computed from the system’s next generation matrix *G* [40]. This takes into account the couplings between the nodes when computing the stability of the disease-free equilibria. The (*i, j*) entry of this matrix corresponds to the expected number of new infections in node *i* caused by an infected individual introduced in node *j*. The next generation matrix, as derived in Appendix A, is given as

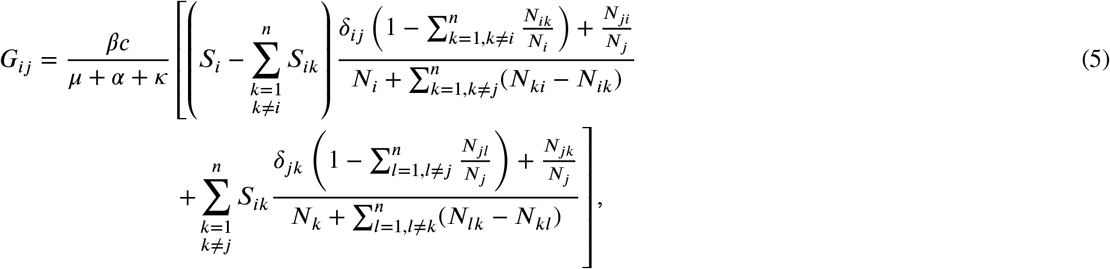

where *δ*_*ij*_ denotes the Kronecker delta: *δ*_*ij*_ = 1 for *i* =*j* and zero otherwise. Then, the effective reproduction number 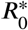 for the networked system is given by the spectral radius *ρ* of the the next generation matrix,

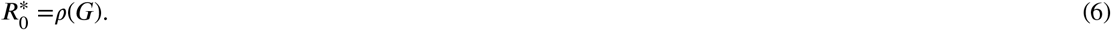

This provides a condition necessary for the stability all disease-free equilibria (*I*_*i*_ = *X*_*i*_ = 0, *S*_*i*_ + *R*_*i*_ + *D*_*i*_ = *N*_*ij*_ for all *i* = 1, …, *n*). It also gives a natural definition for herd immunity in the network, as the networked reproduction number *R*_0_ is now a function of all susceptible populations (cf. the definition of heard immunity for the local dynamics in Sec. 1). All scenarios discussed in the next sections are simulated using our library for modelling epidemics on networks, EpiGraph [24], which is implemented in C++ and freely available on GitHub.

## 3. Case study: Ireland

For the remainder of this discussion paper, we elaborate on the spreading of SARS-CoV-2 in Ireland as a case study. The network model given by Eqs. (4) is informed by publicly available data detailed in the next section.

### 3.1. Network Data

For the Republic of Ireland, each of the 3440 nodes in the network represents an *electoral division* as defined by the central statistics office (CSO), for which small area population statistics are published from the 2016 census [11]. The links in the network are the number of commuters between each electoral division according to CSO data [12]. Similarly, for Northern Ireland, nodes refer to 890 *superoutput areas* according to Northern Ireland Statistics and Research Agency (NISRA) [32]. This spatial resolution corresponds to the local administrative units below the category-3 regions of the *Nomenclature of Territorial Units for* Statistics (NUTS) [19].

### 3.2. 5-Phase Roadmap without Additional Countermeasures

In order to demonstrate the general feasibility of the proposed modelling framework, we consider the 5-phase *roadmap for reopening society & business* by reducing lockdown restrictions as outlined by the Irish Government in May 2020 [16]. Our model attempts to follow the historic evolution of the disease up until phase 2. Then, we allow the model to run under the predicted effects of the remaining lockdown phases. See Tab. 1 for implications on travel and gathering restrictions. Appendix B provides a comparison to reported case numbers. It is important to note that at this stage, we do not consider any reintroduction or roll back to earlier phases during the course of the simulated outbreak. This corresponds to a worst-case scenario and is highly unlikely from a practical point of view, because interventions will be reintroduced, if case numbers rise again.

**Table 1:** Travel and gathering restrictions during the 5 phases of the Irish roadmap.

| Phase | Date | Mobility | Social distancing |
| --- | --- | --- | --- |
| pre lockdown | 22-01-20 | no restrictions | no restrictions |
| school closure | 12-03-20 | no restrictions | recommended reduction |
| initial lockdown | 27-03-20 | travel with 2km radius | essential meetings only |
| 1 | 18-05-20 | travel with 5km radius | essential meetings only |
| 2 | 08-06-20 | travel with 20m radius | maximum of 4 people |
| 3 | 29-06-20 | travel with 20km radius | maximum of 4 people |
| 4 | 20-07-20 | no restrictions | small gatherings |
| 5 | 10-08-20 | no restrictions | large gatherings |
| post lockdown | 31-08-20 | no restrictions | no restrictions |

The parameters are chosen such that the model results match the actual number of deaths in Ireland. See Tab. 2 for details. It is important to note, however, that this model is in the early stages of testing and has not undergone any sensitivity training on the parameters. The effective infection probability *β*_eff_ is computed as *β*_eff_ =*βc* As defined in Eq. (2), the basic reproduction number of the local dynamics [cf. Eqs. (1)] exceeds the critical value of 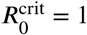 during pre lockdown and after phase 3. Conversely, only initial lockdown, phases 1 and 2 have a *R*_0_ value well below unity and – neglecting network-related effects – exhibit a burn out of an outbreak and self-guided decrease of the number of infected.

**Table 2:** Parameters for the 5 phases of the Irish roadmap. *β*_eff_ and *R*_0_ are computed from the system parameters.

| Phase | $\beta$ | $c$ | $\mu$ | $\kappa$ | $\alpha$ | $\beta_{\text{eff}}$ | $R_0$ |
| --- | --- | --- | --- | --- | --- | --- | --- |
| pre lockdown | 0.37 | 1 | 0.1 | 0.1 | 0.0028 | 0.37 | 1.82 |
| school closure | 0.37 | 0.7 | 0.1 | 0.1 | 0.0028 | 0.26 | 1.28 |
| initial lockdown | 0.37 | 0.4 | 0.1 | 0.1 | 0.0028 | 0.15 | 0.73 |
| 1 | 0.37 | 0.4 | 0.1 | 0.1 | 0.0028 | 0.15 | 0.73 |
| 2 | 0.37 | 0.5 | 0.1 | 0.1 | 0.0028 | 0.19 | 0.91 |
| 3 | 0.37 | 0.6 | 0.1 | 0.1 | 0.0028 | 0.22 | 1.09 |
| 4 | 0.37 | 0.65 | 0.1 | 0.1 | 0.0028 | 0.24 | 1.19 |
| 5 | 0.37 | 0.7 | 0.1 | 0.1 | 0.0028 | 0.26 | 1.28 |
| post lockdown | 0.37 | 0.75 | 0.1 | 0.1 | 0.0028 | 0.28 | 1.37 |

We introduce a small number of diseased individuals to the Dublin area on 22nd January, 2020, that is, 50 days before the initial lockdown, and run the model such that the disease spreads naturally and without control. On the 12th of March, we introduce the initial lockdown measures, which take some time to reach full effect. Individuals’ movement is limited to 2km and this is adhered to with 80% compliance. This level of compliance is supported by preliminary findings of a nation-wide series of phone interviews accounting for essential/justified and non-essential travel [23]. The level of compliance to movement restrictions is kept constant at 80% in the simulations (cf. Fig. 11 for the impact of compliance levels during regional interventions). We account for social distancing and self-isolation (*X* compartment) by reducing the rate of mixing *c* within communities (cf. Tab. 2 for the respective parameter values). During phases 1 and 2/3, the travel radius increased to 5km and 20km, respectively, and we assume the same level of compliance. Travel restrictions are removed in phases 4 and 5 (cf. Tab. 1). Each of the numbered phases have a duration of 21 days. See Appendix D for further details on the impact of movement restrictions on the mobility. In the post-lockdown period, the social mixing parameter is not reset to *c* = 1, but to a lower level of *c* = 0.75 to account for a residual effect of an increased level of awareness compared to pre-lockdown. This is justified, for instance, by ongoing closure of schools and universities and higher levels of home office than at the start of the year.

**Figure 2:**
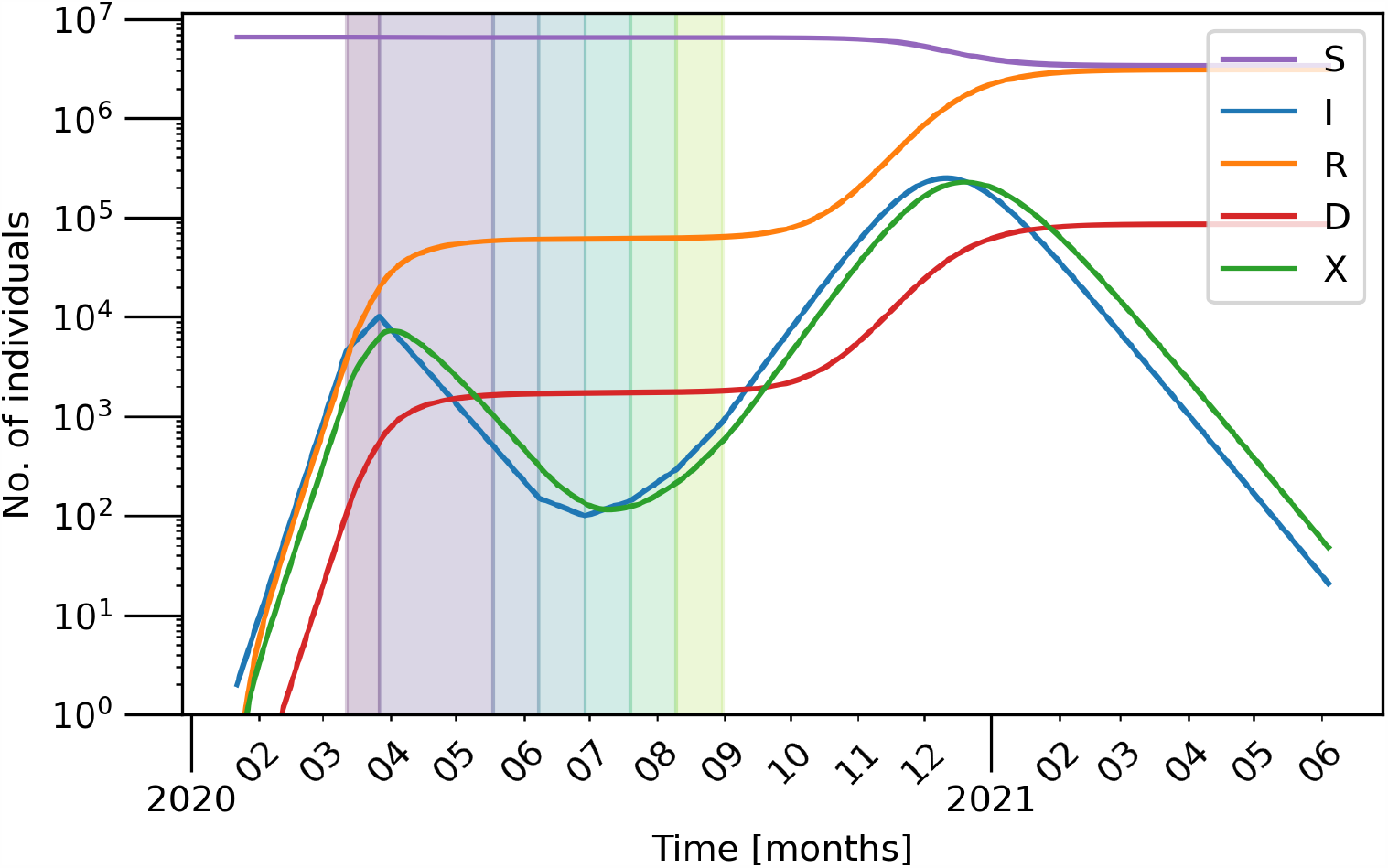
Number of individuals (aggregated over the entire population/network) belonging to each compartment as stated in the legend over time with a log scale on the y-axis. Parameters as in Tab. 2 and an 80% compliance to movement restrictions. Each successive lockdown phase is indicated by the differently coloured shaded regions on the plot. After the initial lockdown (violet), phases 1 – 5 last 3 weeks each.

**Figure 3:**
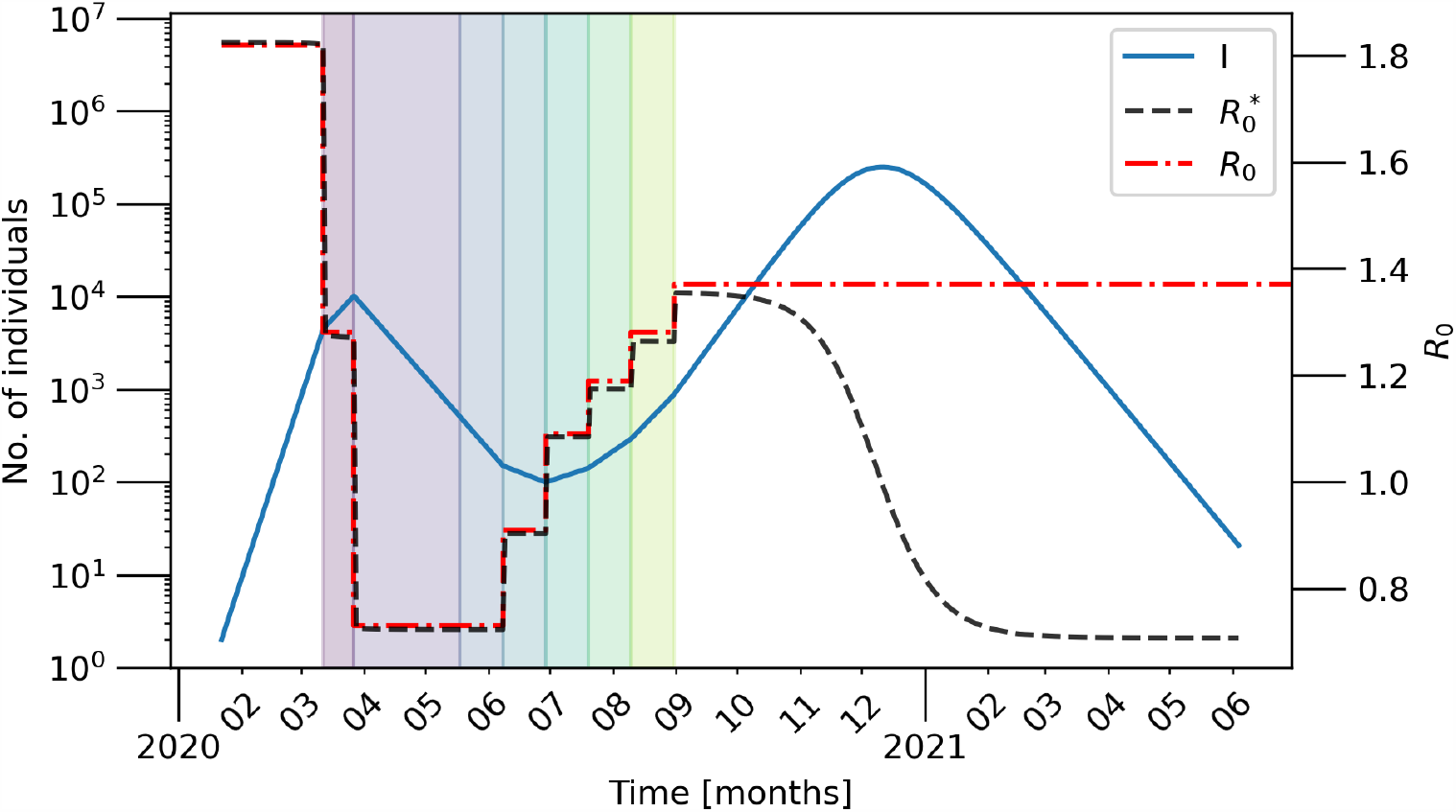
The basic and effective reproduction numbers *R*_0_ (black dashed) and 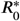 (red dash-dotted) over time as calculated from the basic reproduction number and spectral radius of the next generation matrix given by Eq. (5), respectively, together with the number of infected (blue solid) in the network in the doomsday scenario as shown in Fig. 2.

**Figure 4:**
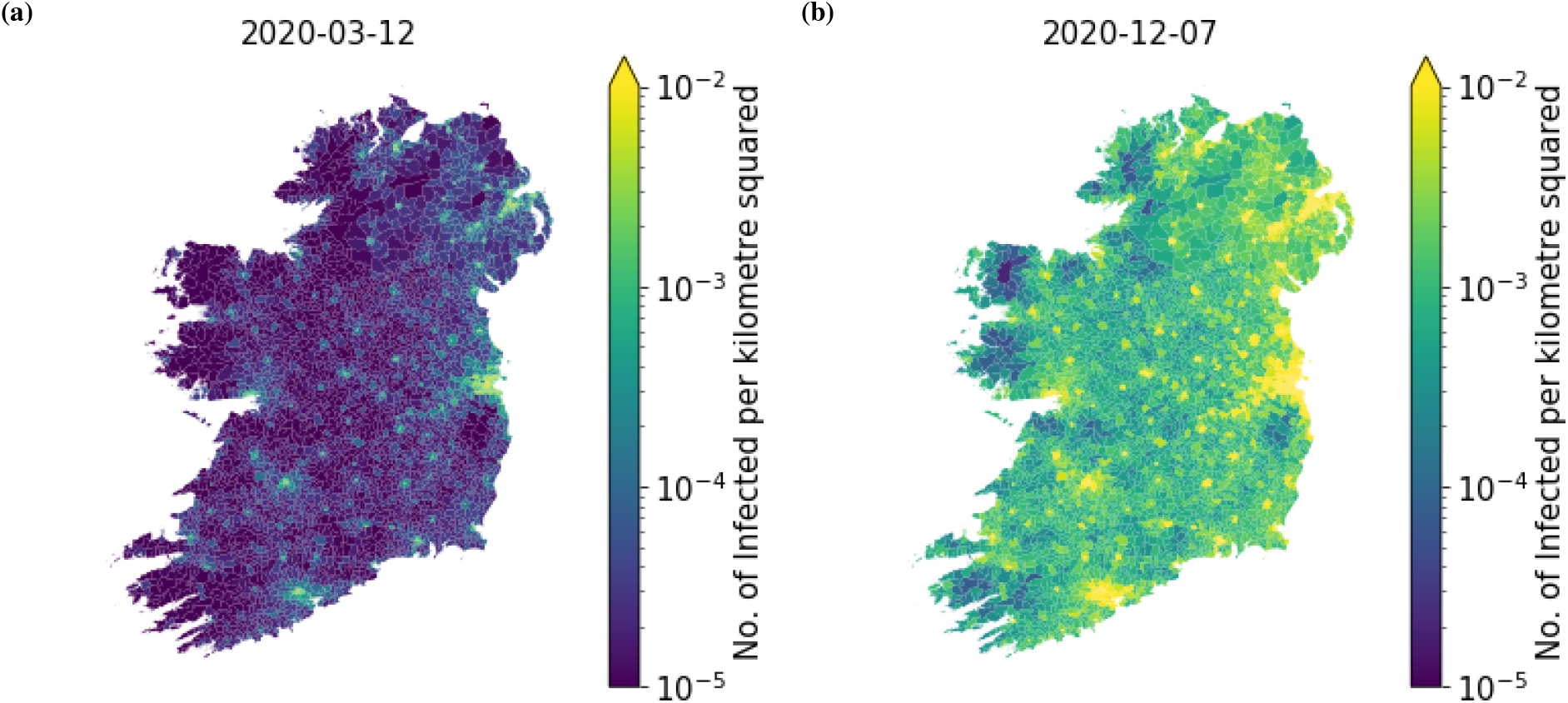
The spatial distribution of infected individuals per km^2^ around the two distinct peaks shown in Fig. 2 at (a) Day 50 (b) Day 320. Parameters as in Tab. 2.

**Figure 5:**
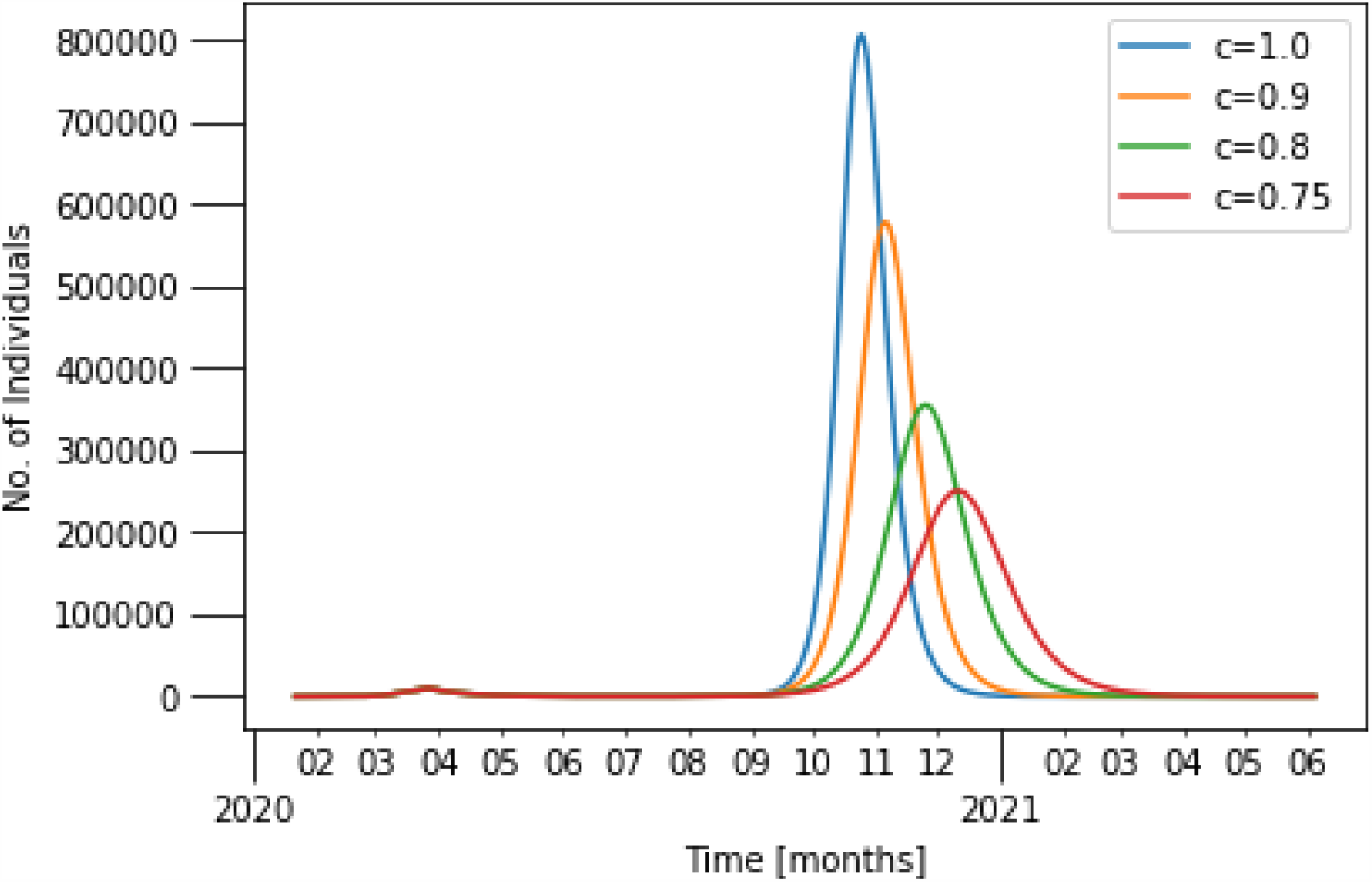
Evolution of the aggregated, infected compartment *I* for varied post-lockdown conditions. The blue curve corresponds to the parameters given in Tab. 2 (cf. Fig. 2 for a semi-logarithmic scale). The other curves vary the parameter *c* in the post-lockdown phase as stated in the legend: *c* = 0.9, *c* = 0.8 and *c* = 0.75. Other parameters are as given in Tab. 2.

**Figure 6:**
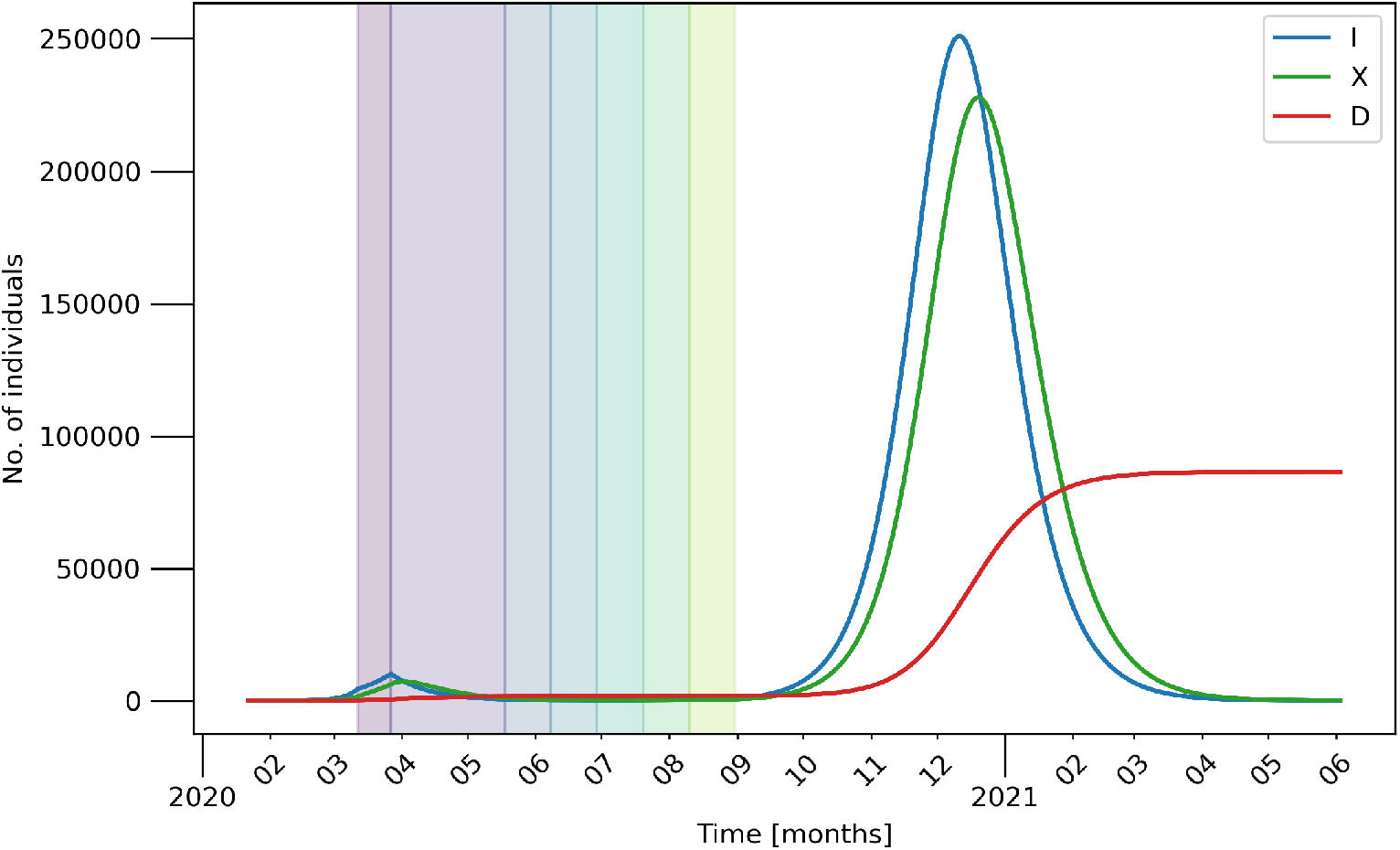
Time series for 10 random initial conditions, where each curve is superimposed. Note that it is impossible to visually distinguish between the different time series for time each respective compartment as each curve lines up almost perfectly with all the others. The social mixing parameter is chosen as *c* = 0.75. Other parameters as in Tab.2.

**Figure 7:**
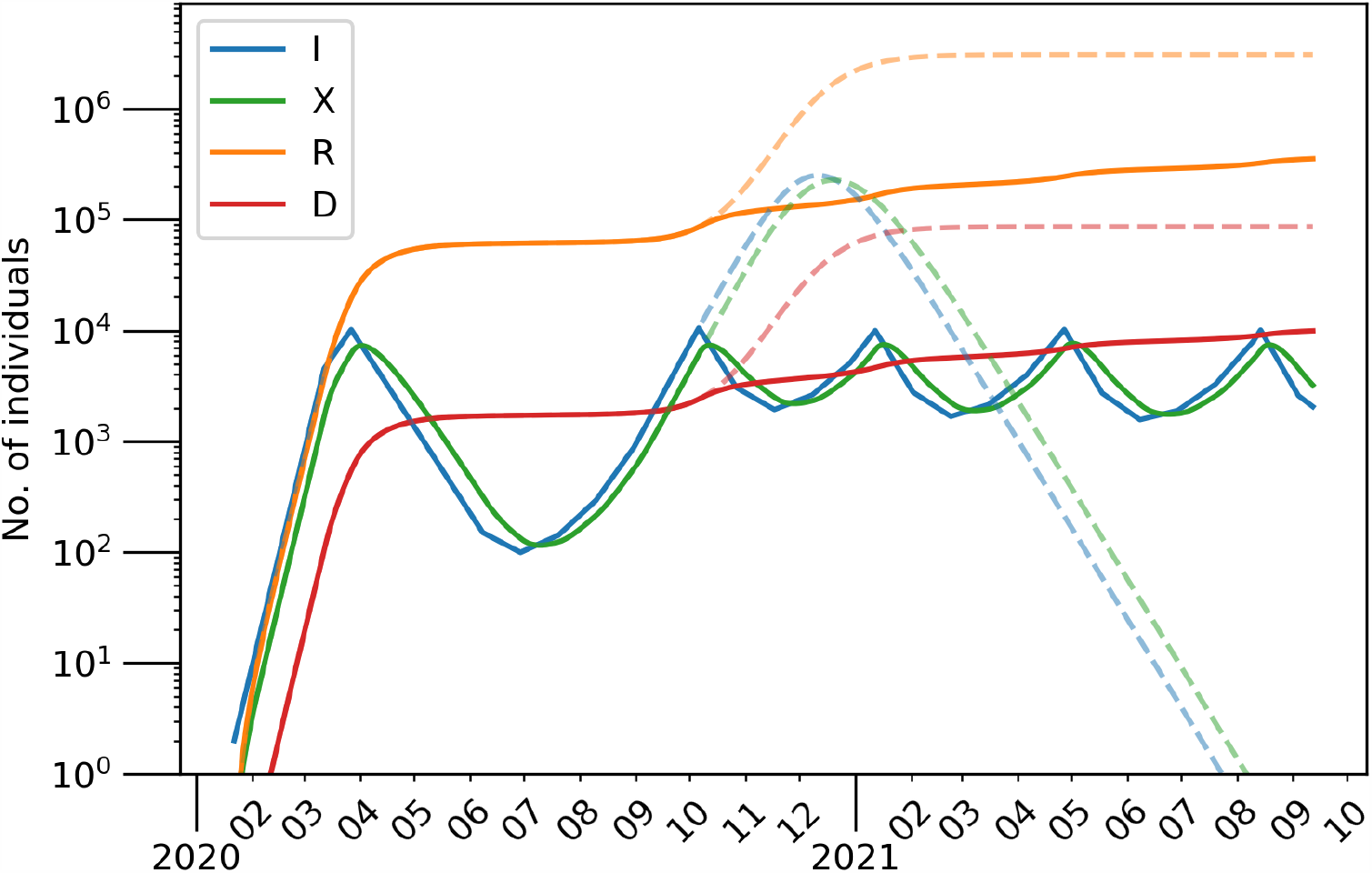
The number of individuals belonging to each compartment over time (using the parameters in Tab. 2) under a dynamic lockdown strategy taking *I*_th_ = 10^4^. The dashed lines are the equivalent model without the dynamic lockdown strategy shown in Fig. 2.

**Figure 8:**
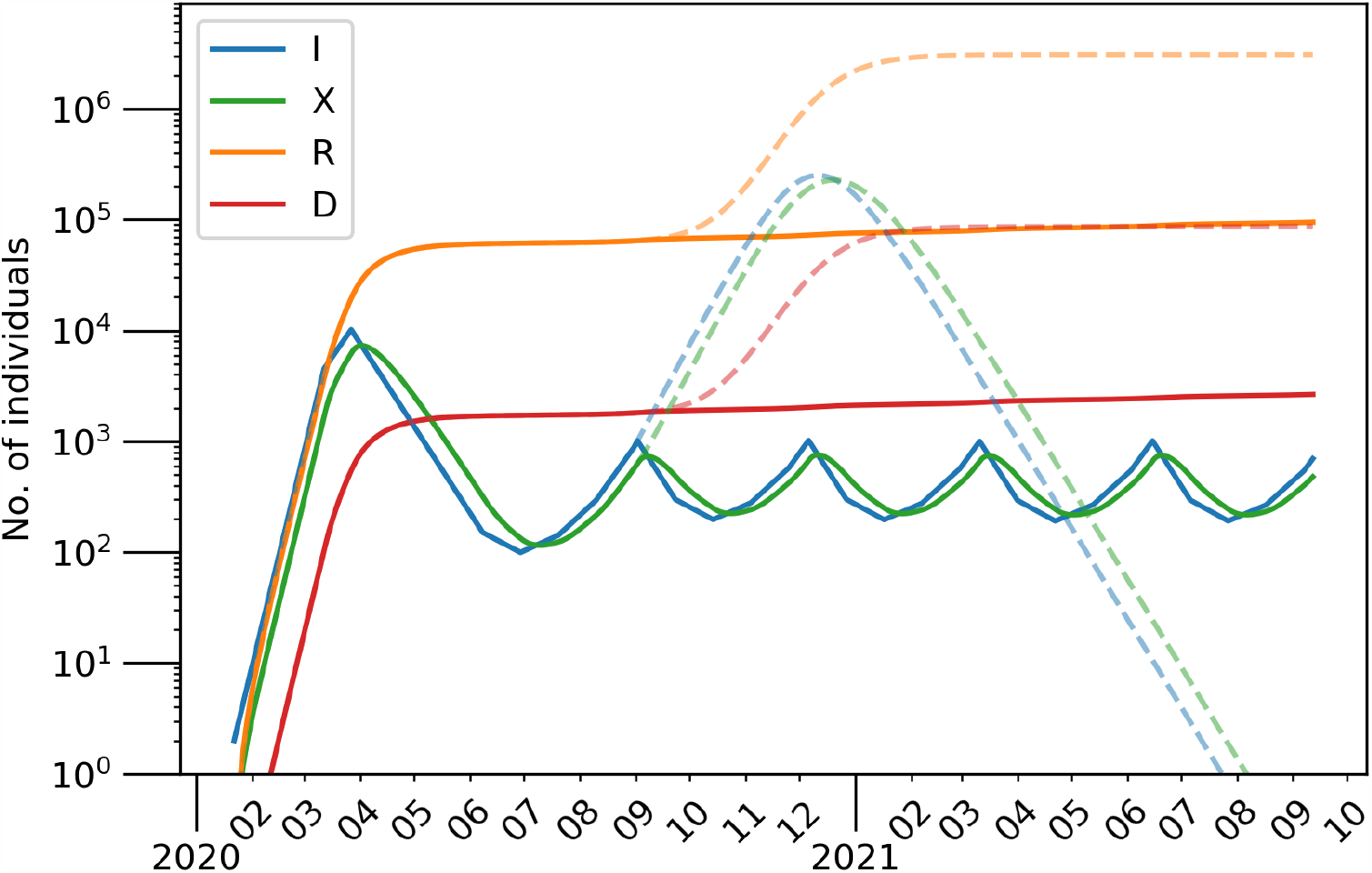
The number of individuals belonging to each compartment over time (using the parameters in Tab. 2) under a dynamic lockdown strategy taking *I*_th_ = 10^3^. The dashed lines are the equivalent model without the dynamic lockdown strategy shown in Fig. 2.

**Figure 9:**
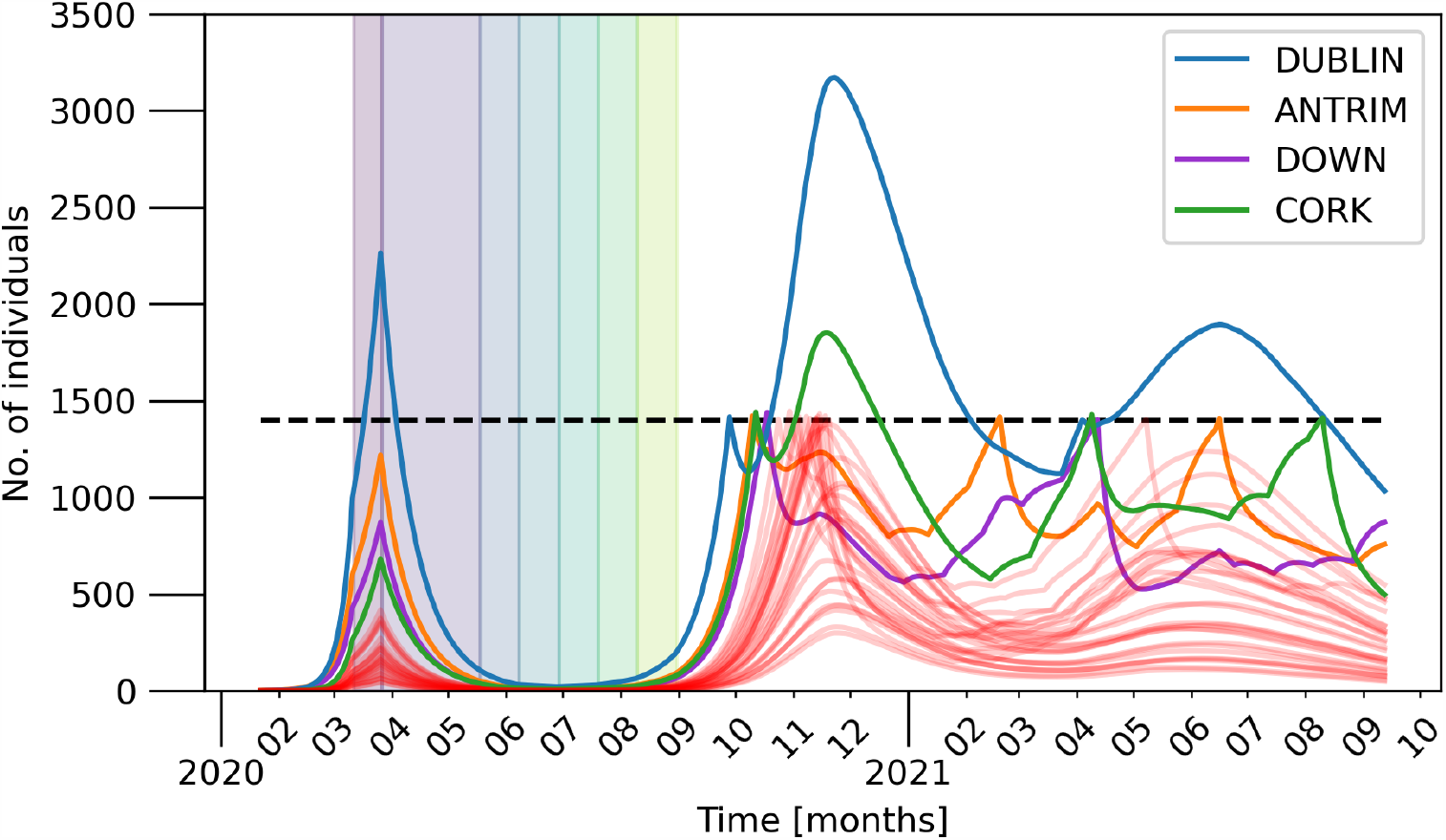
Number of individuals (aggregated over counties) belonging to the *I* compartment. Parameters as in Tab. 2. Each successive lockdown phase is indicated by the differently colored shaded regions on the plot up until the lockdown phases for each county become desynchronised. After the initial lockdown (violet), phases 1 – 5 last 3 weeks each. The four largest counties by population are explicitly denoted in the legend.

**Figure 10:**
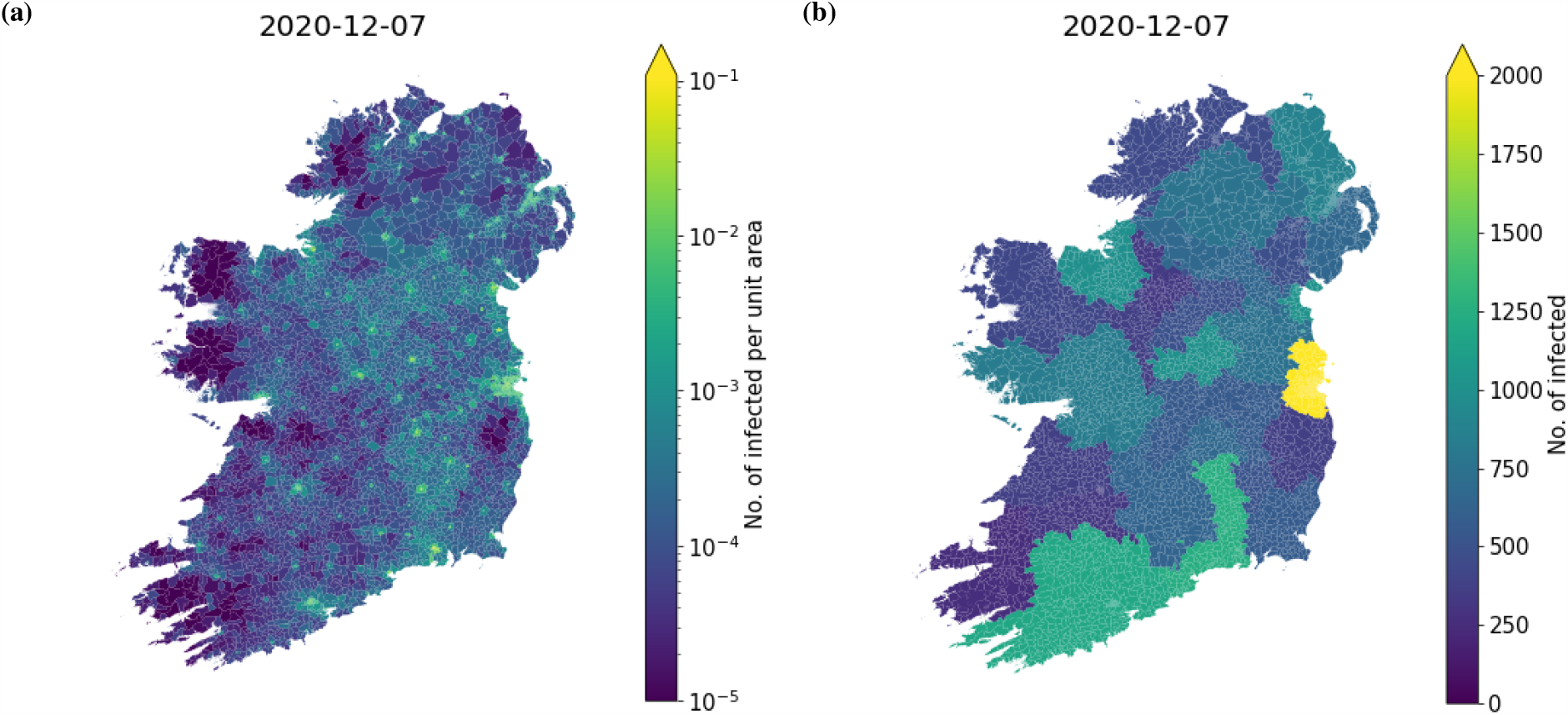
The geographic distribution of infected individuals in the simulation shown by the time-series in Fig. 9 on the 7th December, 2020. Panel (a) shows the number of infected per km^2^ in each electoral division and superoutput area. Panel (b) shows the number of infected in each county.

**Figure 11:**
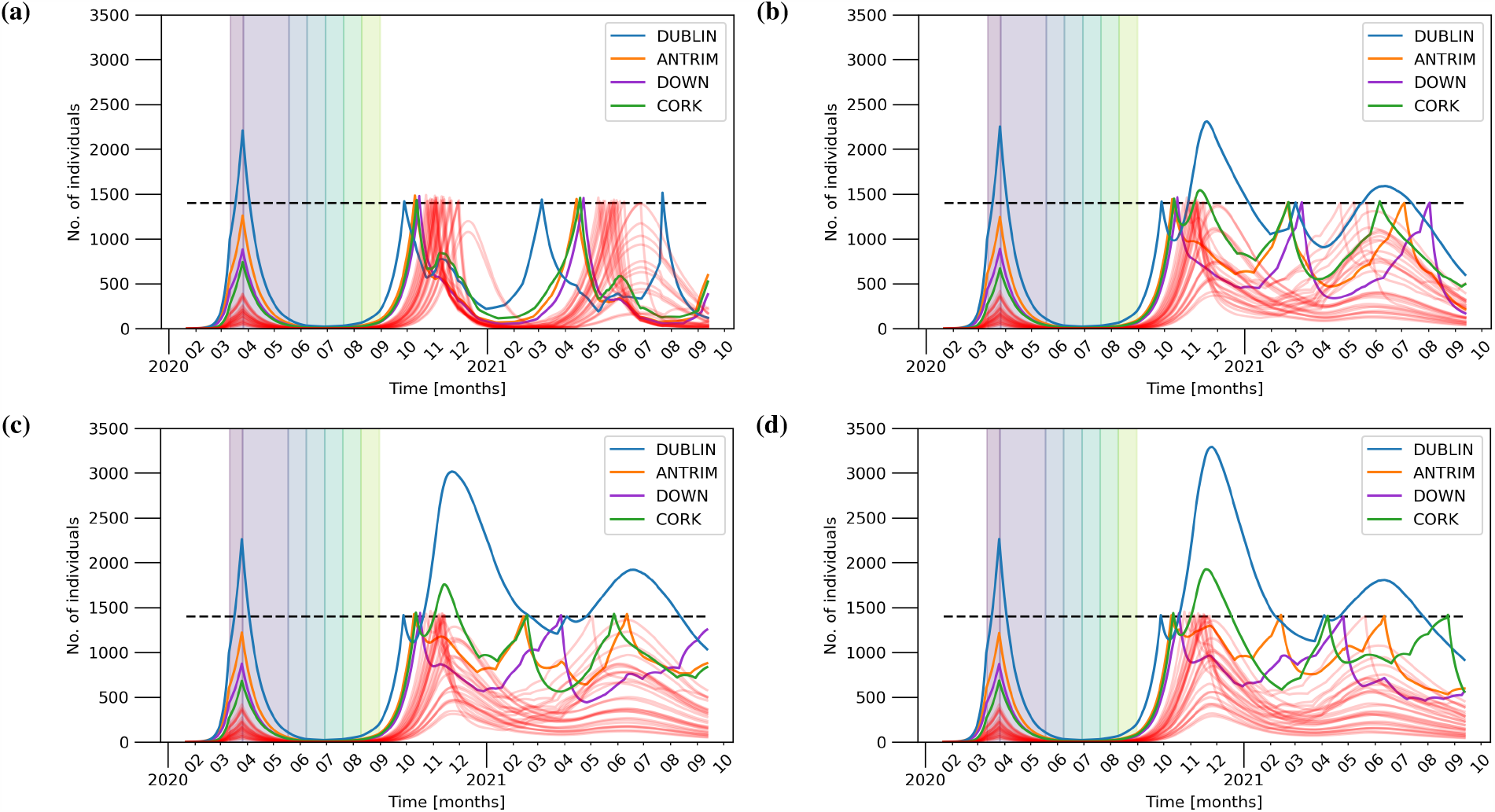
Number of individuals (aggregated over counties) belonging to the *I* compartment for compliance of 100%, 99%, 90%, and 70% in panels (a), (b), (c), and (d) respectively. Other parameters as in Tab. 2. Each successive lockdown phase is indicated by the differently colored shaded regions on the plot up until the lockdown phases for each county become desynchronised. After the initial lockdown (violet), phases 1 – 5 last 3 weeks each. The four largest counties by population are explicitly denoted in the legend.

Figures 2 depicts the time series of all five compartments – aggregated over all nodes – in semi-logarithmic scale. One can see two distinct peaks: (i) a small peak that occurs before the lockdown and (ii) a second peak that occurs once all lockdown measures are lifted. It is clear that unless some of the measure are maintained, then the lockdown will just have the effect of delaying the epidemic and result in a second wave with a prevalence that – in this unlikely worst-case scenario – is 100 times higher than the first one and exceeds any realistic capacity of the health-case system. The number of infected eventually reaches zero due to a depletion of the pool of susceptibles. Following the geometric analysis of the local dynamics (cf. Sec. 1), the trajectory lands at one of disease-free equilibria *I*_e_ = 0. A change in parameters, e.g., rate of mixing *c*, or a rescaling of the population to the number of remaining susceptibles can render this state unstable again. Then, a perturbation to the networked system, for instance, introduction or importation from outside, has the potential to trigger an additional outbreak.

For the simulation run given in Fig. 2 we compute the next generation matrix (5) at each time-step and find its spectral radius giving rise to the network’s effective basic reproduction number 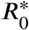, which is not only a function of the model parameters but also the number of susceptibles. For details, see Appendix A. 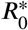 is plotted as black dashed curve in Fig. 3. For comparison, the local/simplistic value of *R*_0_ given by Eq. (2) is shown as (dash-dotted) red curve. The value of 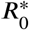 shows the potential for the disease to spread at each phase in the simulation. From Fig. 3 we can see clearly that when 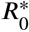 falls below unity, the number of infected also begins to fall. We also note the point of herd immunity is reached is in when the number of susceptibles crossed the point where the 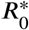 goes below 1.

As stated above, the parameters are chosen to match the actual number of deaths in Ireland. This, in turn, results in a larger first peak compared to the reported numbers of confirmed cases on the 12th of March ranging from 27 according to [21] to 70 according to [44] (cf. Fig. 13 in Appendix B for a detailed comparison to reported data). The simulated results, however, are not meant to reflect this range of confirmed cases, but following the logic of the model, refer to all infected individuals including asymptomatic, non-diagnosed and untested.

**Figure 12:**
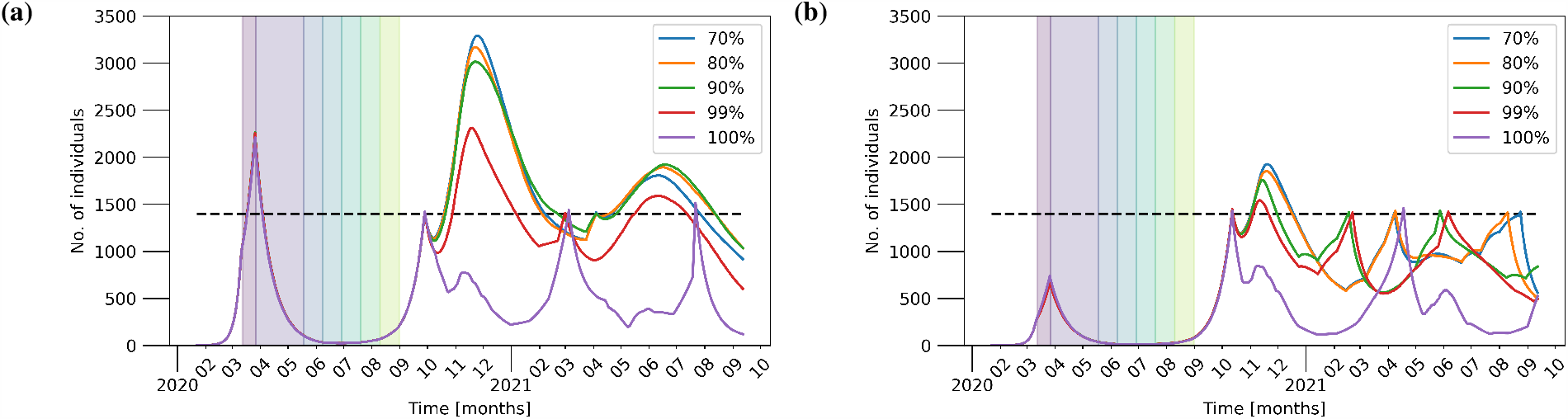
The time-series for the evolution of the *I* compartment in (a) Dublin and (b) Cork extracted from Figs. 11 and 9 for different values of compliance: 70% (blue), 80%(orange), 90% (green), 99% (red), and 100% (blue).

**Figure 13:**
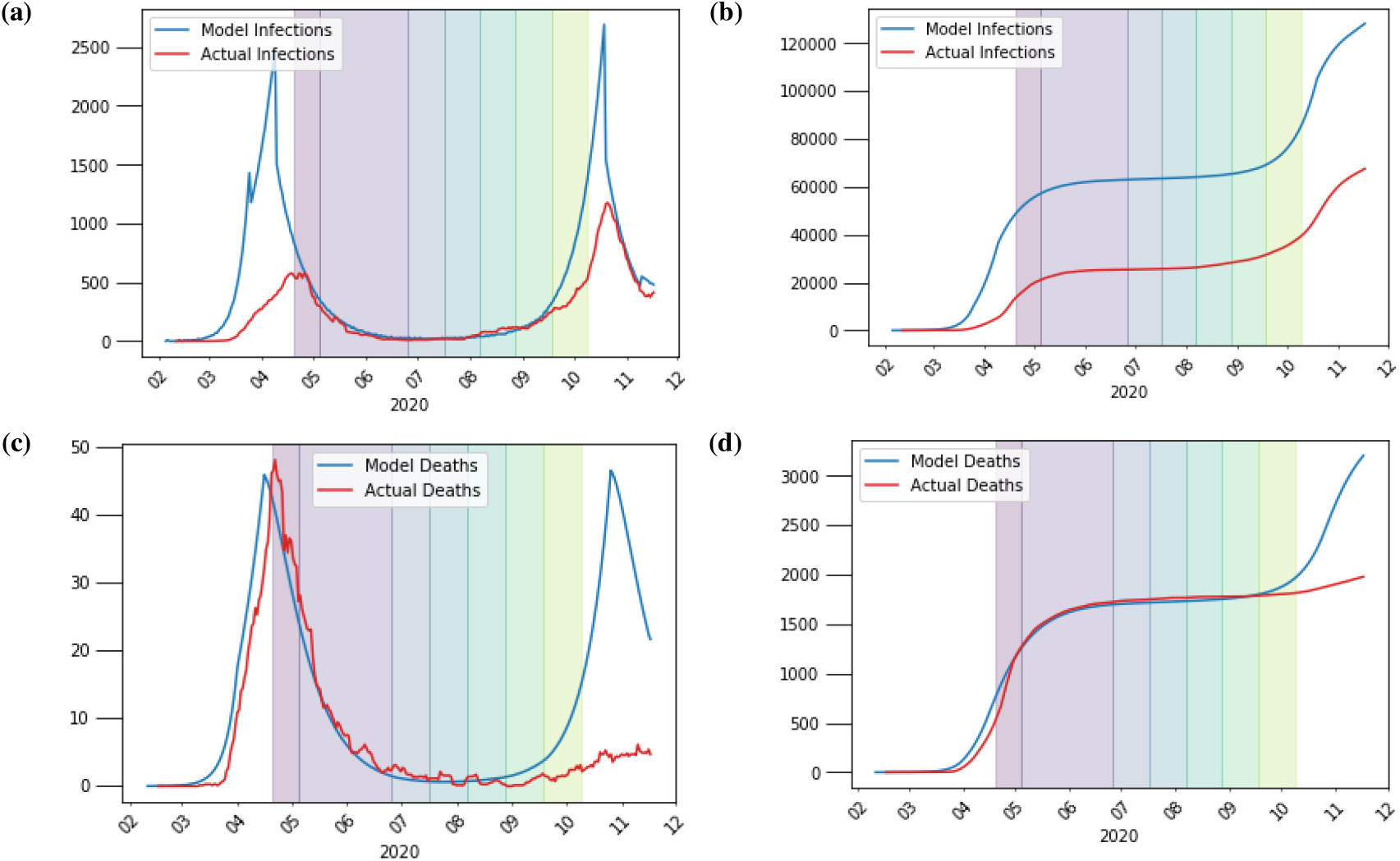
Comparison of the countrywide dynamic lockdown scenario with threshold of infection *I*_*thresh*_= 10000 (blue) to actual figures (red) for reported daily and cumulative infections (panels 13a and 13b) and daily and cumulative deaths (panels 13c and 13d) from the Health Protection Surveillance Centre (HPSC) in Ireland. Other parameters are as given in Tab. 2.

Figures 4a and 4b show spatially resolved snapshots of the prevalence of the disease per km^2^ at times *t* = 50d (March 12) and *t* = 320d (December 7), respectively (cf. [22] for a map based on data of reported, confirmed cases). Figure 4a corresponds to the disease prevalence at the first peak before the lockdown. The disease remains in the East of the country for the most part with the exception of big towns and cities. In contrast, at *t* = 320d (Fig. 4b), the outbreak covers the entire country and affects every electoral division. See Fig. 2 for a full time series of the aggregated variables.

Figure 5 shows the effect of different levels of awareness during the post-lockdown phase, which is modelled by different values for the mixing parameter *c*. For convenience, the plots only depict the time series of the aggregated infected compartment *I* in a linear scale. A reduction to 70% of the original probability of infection significantly leads to a significantly smaller and delayed second peak (red curve), while the basic reproduction number of the local dynamics remains above unity (cf. Tab. 2).

In order to show how the heavy-tail distribution of travel across the network causes a lack of sensitivity in initial conditions, we run the model for a number of random initial seeds where for each run only a single individual is placed in one electoral division or superoutput area at the start of the simulation. The resulting time series are shown in Fig. 6.

One can see that both amplitude and timing of the large, second peak are stable and it is impossible to visually differentiate between any of the curves for different initial conditions. In the presented 4 cases, the initial location is selected uniformly at random from all network nodes. We note that deliberately selecting urban or rural locations will have no qualitative impact. To further corroborate the results, initial infections of a significant, finite fraction of the population will be explored. This might account for a considerable amount of unreported cases at the start of the recording of SARS-CoV-2 cases.

### 3.3 Dynamic Interventions

Finally, we discuss the likely scenario of a flexible lockdown policy that monitors the prevalence of the disease within the island and aims to keep it below the capacity of the health-care system. As a simple measure, we define some threshold *I*_th_ as the maximum number of infected that can be present before we go back into lockdown and go through earlier phases again. Such a procedure corresponds to dynamic [14, 27, 28] or active [4] interventions. Specifically, we simulate until phase 3 as before to maintain the agreement with reported cases. Afterwards, if the condition *I*_th_ > Σ_*i*_ *I*_*i*_ is met, the system goes back to phase 1 and then progresses from there until the condition is met again.

#### Island Interventions

In detail, we consider two scenarios with different threshold parameters, which are inspired by the reported, maximum number of hospitalized cases during the first wave in April 2020 [22]: (i) *I*_th_ = 10^4^, as shown in Fig. 7 and (ii) *I*_th_ = 10^3^ as depicted in Fig. 8. In both figures, the worst-case scenario of Fig. 2 is added as dashed curves. Note that the curves of recovered *R* (orange) and deaths *D* (red) are only increasing at a low rate. For the case of *I*_th_ = 10^4^, for instance, these values arrive at 10^5^ after two years and thus, way below any effect of herd immunity.

Under the assumptions the considered network model, it would take many years for a significant depletion of the pool of susceptibles. Extrapolating from an extended run of the model similar to Fig. 7, we find that it would take roughly 27 years for the number of recovered to reach half the population, leading to a total number of approximately 92,000 deaths over the course of many waves. Even with a large threshold *I*_th_ = 10^4^ for the maximum number of infected cases, the feasibility of such a strategy is questionable due to the time it would take to reach herd immunity of on the island.

The re-introduction of lockdown measures to account for rising numbers of infected can change the stability of the disease-free equilibria *I*_e_ = 0. However, as soon as the restrictions are relaxed, e.g., during phases 3 to 5, an unstable family of these equilibria re-emerges and the outbreak pattern repeats. Suppressing the occurrence of unstable disease-free equilibria requires other forms of interventions. One could consider, for example, a fishing-type extension of the model [3, 6, 15]. This will basically turn the rate *κ* in the local dynamics (1) into a function of the number of infected, e.g., 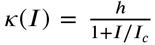 with a consumption/fishing rate *h* (here: rate of isolation) and a reference capacity *I*_*c*_ for, *κ* (*I*) = *h*/2. This *I* dependence will account for contact-tracing effects, which are especially effective for low numbers of infected. As a consequence, a high rate of isolation will be able to push the basic reproduction number [cf. Eq. (2) or Eq. (6)] below the critical value of 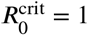 and change the stability of the equilibria *I*_e_ = 0.

#### Regional Interventions

Next, we consider another scenario that accounts for regional lockdowns at the level of counties. In our dynamic intervention model this corresponds to each county having its own 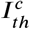. Thus, once the county surpasses this threshold, another lockdown is triggered sending the county back to phase 1^*th*^. When lockdown phases differ between counties, we assume that the travel restrictions in and out are followed with the compliance of the county being travelled to. When we run the simulations using the regional interventions, the model operates at the level of electoral divisions and superoutput areas but their travel and interaction dynamics are consistent across counties. The parameters used for each phase are the same as those given in Tabs. 2 and 1. For this run we took the maximum number of infected per county 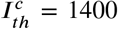 and as usual the model is allowed to run without intervention until phase 3 to match the historic number of deaths.

From Fig. 9 we see some interesting dynamics when we view the evolution of the number of infected in each county. After Dublin re-enters phase 1 due to dynamic lockdown after only a few days the numbers start rising again. This is not due to its own dynamics, but due to its interaction with neighbouring counties, which are not yet in lockdown. It is only once all the other counties enter phase 1 lockdown do we see the numbers in Dublin and Cork begin to drop. We conclude that treating such counties in isolation is not a good strategy due the strong coupling between each county that is not eliminated when some counties remain free to travel even only within their own borders. Figure 10 shows the geographic distribution of the disease during the largest peak of Dublin seen in Fig. 9 at the level of electoral divisions and superoutput areas in panel (a) and at the level of counties in panel (b).

Next, we investigate the impact of compliance to movement restrictions. Figure 11 depicts the time series of the *I* compartment for 100%, 99%, 90%, and 70% compliance (cf. Fig. 9 for the case of 80% compliance). One can see that already a small amount of non-compliance results in outbreaks with numbers above the threshold. For example, 99% compliance shown in panel (b) triggers numbers of infected as high as during the first wave in March/April 2020.

Figure 12 shows the time evolution of infected aggregated over both Dublin and Cork county for different levels of compliance (cf. Figs. 9 and 11). These examples show that lower compliance yields higher peaks after the introduction of regional interventions.

## 4. Conclusion

We have proposed a dynamical network model to study the spreading of SARS-CoV-2 that accounts for travel or commuting between nodes of the network. As a case study, we have presented results from numerical simulations for Ireland, where the model is informed from publicly available data on geographical regions, population, and mobility in both the Republic of Ireland and Northern Ireland. The network model is flexible in terms of spatial granularity and parameter selection. We have focused on the governmental 5-phase roadmap for reopening society & business and considered different initial conditions, parameters, and mobility data including realisitic levels of adherence to movement restrictions. We have observed that the roadmap will lead to a decline of case numbers. However, once the restrictions are lifted, the numbers will rise again, even if an increased level of awareness is considered. This is in excellent agreement with reported case numbers (cf. Appendix B).

Finally, we have elaborated on a procedure of dynamic interventions that re-introduce lockdown measures, if a certain threshold of infection is reached. Following such a protocol, the overall number of infected cases can be controlled and safely kept below a prescribed threshold, which can be selected according to the capacity of the healthcare system. We have considered that containment measures would be re-introduced on a country-wide level, but a more regional approach (county level) appears feasible as well. It is likely that in the long term, the disease will be contained best by vaccination and not by the development of natural herd immunity [2]. While we have not considered the effects of vaccination in this paper, in our model naturally acquired herd immunity, or population resistance, will develop slowly over a long period of time. This is true even with our assumption that infection confers long-lasting resistance, and it is far from clear that this actually the case. Therefore the model supports the view that effective herd immunity will best be obtained with widespread immunisation [2] although factors such the duration of immunity, efficacy in different age demographics or ethnic groups, likely vaccination uptake, effects of different vaccines, as well as efficacy against novel variants are poorly understood. Our model could be used to study these variables but this is outside the scope of this paper.

We hope that the results presented in this discussion paper will stimulate scientific and public discussion beyond the island of Ireland about the purpose and effect of lockdown measures in the months ahead (cf. Appendix C). These are plausible scenarios given the time frame of vaccination campaigns that have just begun.

## Data Availability

All data sets used in the present study are publicly available and links are provided in the list of references. The code is available via GitHub.

https://github.com/rory-humphries/EpiGraph-cpp

## Acknowledgements

The work is supported by Health Research Board Ireland in the framework of the COVID19 rapid response programme (grant number: COV19-2020-117). We are grateful to Andreas Amann, Patricia Kearney, Gerry Killeen, and Ivan Perry for helpful discussions.

## Authors’ contributions

RH, KM, SW, MOR, and PH designed the study. SW and RH developed the theory. RH and MS performed the numerical simulations and analyzed the data. All authors discussed the results and contributed to the writing of the article. All authors read and approved the final version of the manuscript.

## A. Next Generation Matrix

In this appendix, we provide a detailed derivation of the next generation matrix given Eq. (5). We start by splitting the networked SIXRD equations (4) into the parts that cause new infections and all other changes. We shall denote the rate of appearance of new infections at node *i* in compartment *U* by ℱ_*U,i*_ (*x*). Similarly, let 𝒱_*U,i*_ (*x*) be the rate of change of individuals in compartment *U* and node *i* by all other means such that 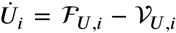. This yields the following equations:

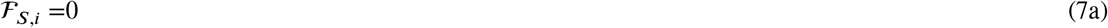

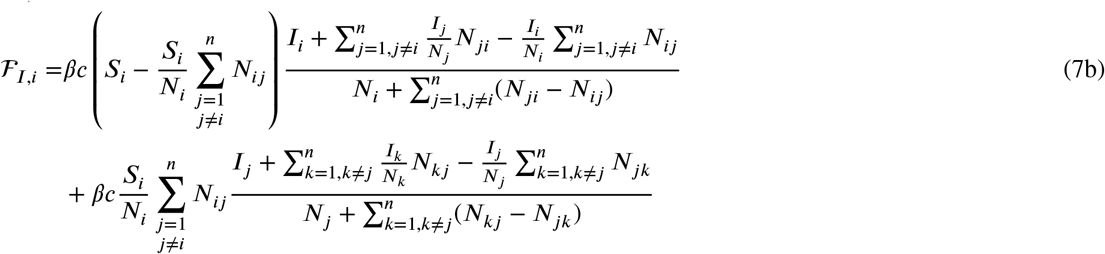

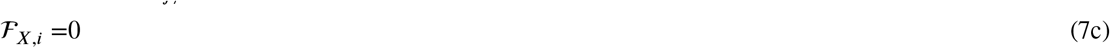

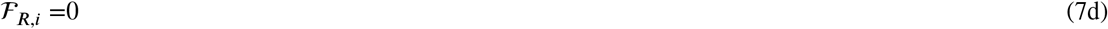

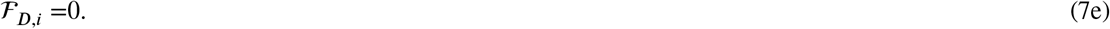

It is an important note that the movement of individuals form *I* to *X* does not count as new infections. Details on the impact of the travel statistics during the different phases of the roadmap can be found in Appendix D.

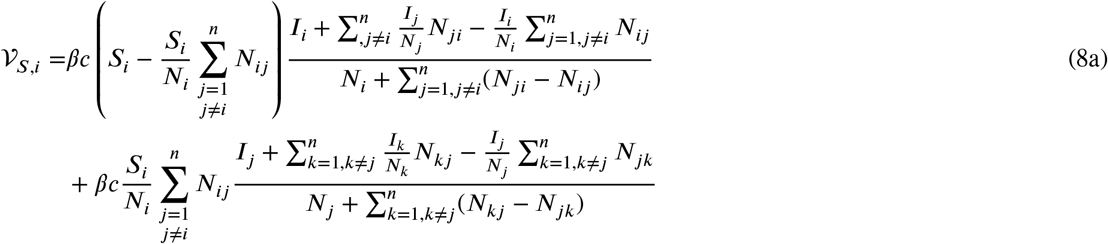

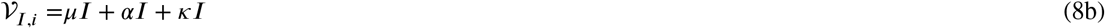

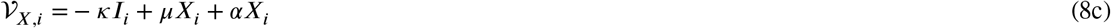

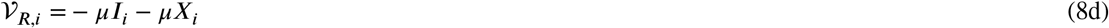

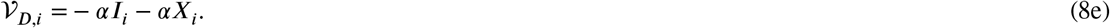

For the above equations, we replace the coupling variables with their actual values, i.e. we take 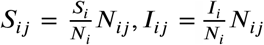, … etc. Writing these equations in vector form we obtain

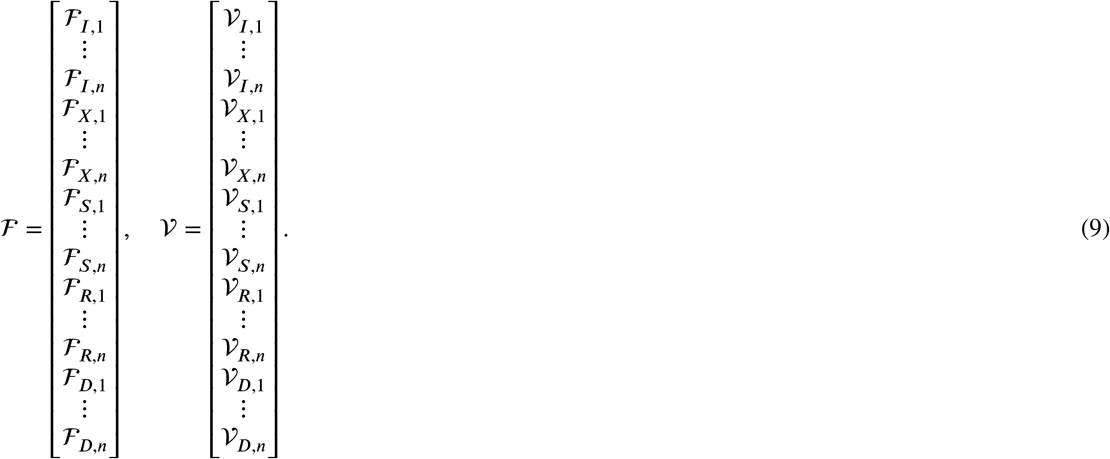

The disease-free equilibria are all solutions that satisfy

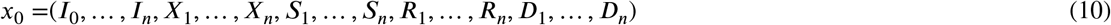

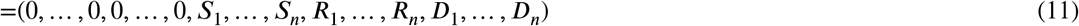

Taking the derivatives of F and V at the disease-free equilibria, we find that

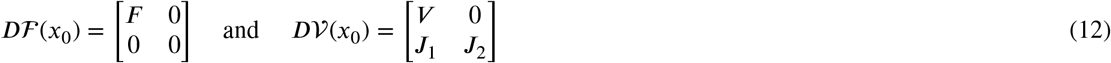

as shown in [40]. We obtain

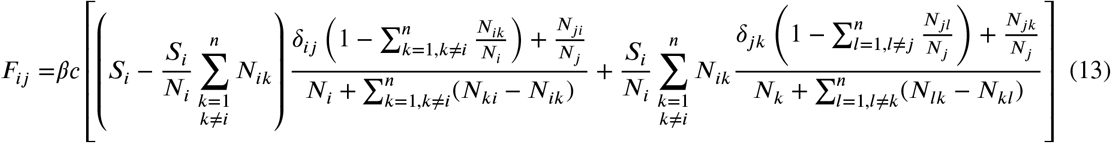

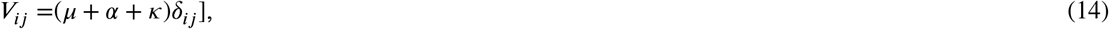

where *δ*_*i,j*_ denotes the Kronecker delta: *δ*_*i,j*_ = 1 for *i* =*j* and zero otherwise. Finally, following [40], the next generation matrix is given by

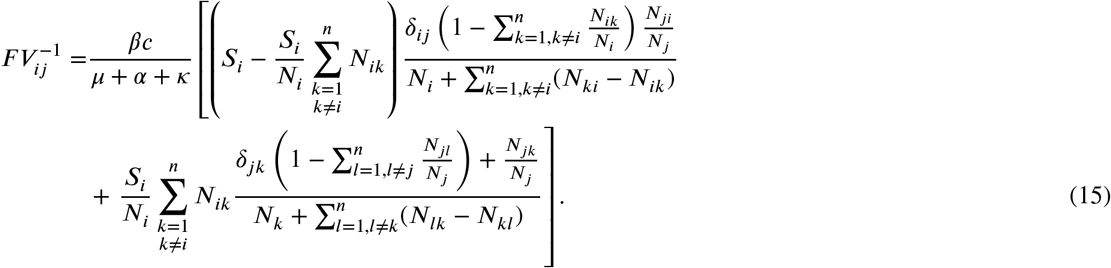

## B. Comparison to Real-World Data

Figure 13 shows a comparison of model results for a countrywide dynamic lockdown scenario with daily and cumulative figures for reported deaths and infections from the Health Surveillance Centre (HPSC). Following the initial lockdown and phased reopening as outlined in Tab. 1, the threshold of infection was chosen to be 10000 in these simulations, before a national lockdown was reintroduced. This figure is adjustable based on current public health guidelines. Panels 13a and 13b compare the model to daily and cumulative reported figures for infections, respectively. Moreover, panels 13c and 13d compare the model results to daily and cumulative figures for deaths. The blue curves correspond to the model results, and the red curves refer to the data reported by HPSC. A 14-day moving average has been applied to data from the HPSC. Note that the infected compartment in the model does not distinguish between confirmed/undetected symptomatic/asymptomatic cases. This explains the larger number of infections in the model in which we see a clear qualitative agreement in the number of infections over time. The number of deaths, however, is more reliable and the model reproduces the actual numbers with large agreement. Overall, we find that the model results compare favourably to these real-world data.

## C. Projected Post-Christmas Lockdown

As a final scenario, we finish with a projection on the a possible post-Christmas projection during which we expect to see the third wave of lockdown. Figures 14 considers the current period of the second wave and the corresponding lockdown with level-5 movement restrictions in the Republic of Ireland. After a partial lifting of these measures scheduled for 1st December 2020, we find that the case numbers will rise again. Note that the numbers remain significantly higher than during summer 2020. This is illustrated by the maps in Fig. 15, which compares the situation by the end of June and 1st December. We project that the numbers will reach a high level similar to the introduction of the current lockdown by late-January 2021 and thus will require another lockdown of similar duration.

**Figure 14:**
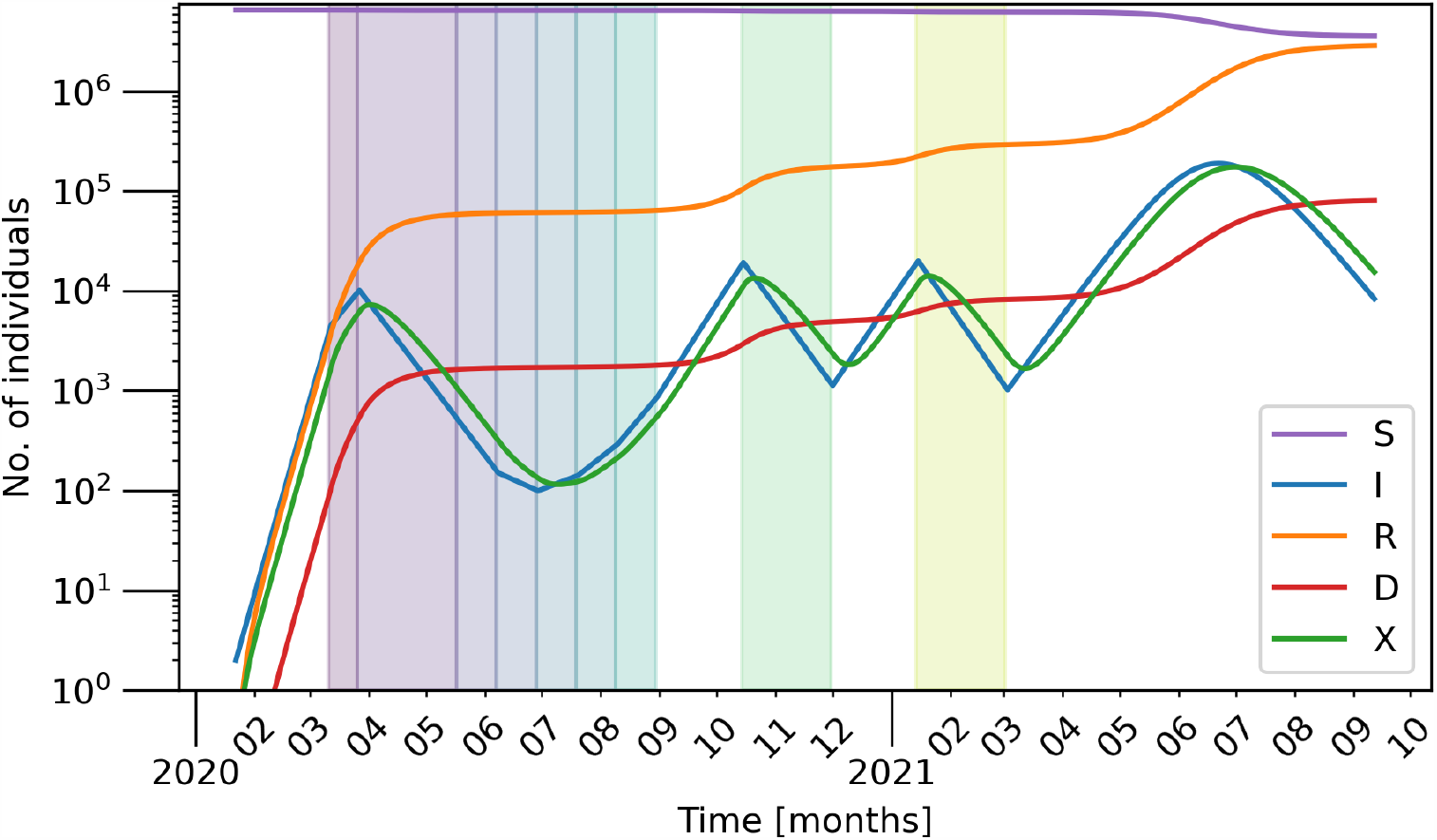
Simulating the evolution of the disease such that the historic deaths are matched up until level 5 and projected form then on. There are two subsequent periods of lockdown: the current lockdown that started mid-November and is expected to lift 1st December. The next lockdown, which occurs in January 2021, is projected based on the number of the numbers of the November lockdown. Other parameters are as given in Tab. 2.

**Figure 15:**
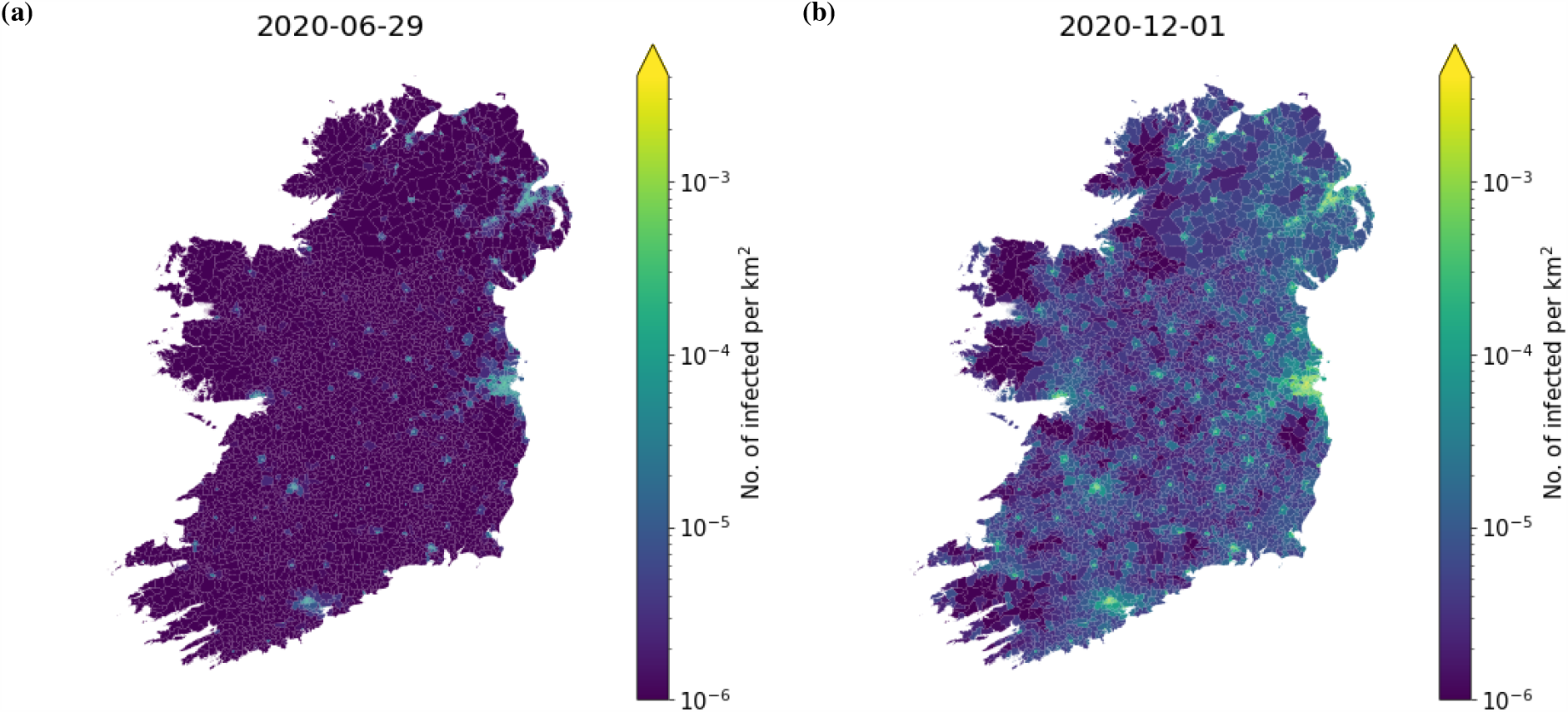
The geographic distribution of infected individuals per km^2^ in each electoral division and superoutput area in the simulation shown by the time-series in Fig. 14 Panel (a) shows the geographic distribution during the lowest incidence of infected during the summer and panel (b) shows the geographic distribution at the end of the lockdown projected to end the 1^st^ December 2020. Other parameters are as given in Tab. 2.

## D. Movement data

In this appendix, we demonstrate how travel restrictions affect the movement of individuals in the simulations. We extracted the number of travelling individuals *N*_*ij*_ between locations *i* and *j* from CSO data and generated a probability distribution of travelling from one electoral division/super-output area to another as a function of population density and distance. Once the travel restrictions are put in-place, the probabilities of travelling to areas outside this distance are scaled by the compliance factor making individuals less likely to travel there. Each *N*_*ij*_ was then generated from these probability distributions inferred from real commuting data within the country.

The difference in these movement patterns can be seen in Fig. 16 where the cumulative distribution of travel disances is plotted for each level of travel restrictions (2km, 5km, and 20km) and the original data prior to the first lockdown period, which are used during periods with no travel restrictions (cf. Tab. 1). The inset shows the original disttribution for the full range in a semi-logarithmic scale. The kinks in the distribution of the restricted movement occur at the respective radii. Larger distances still occur due to non-perfect compliance (here: 80%).

**Figure 16:**
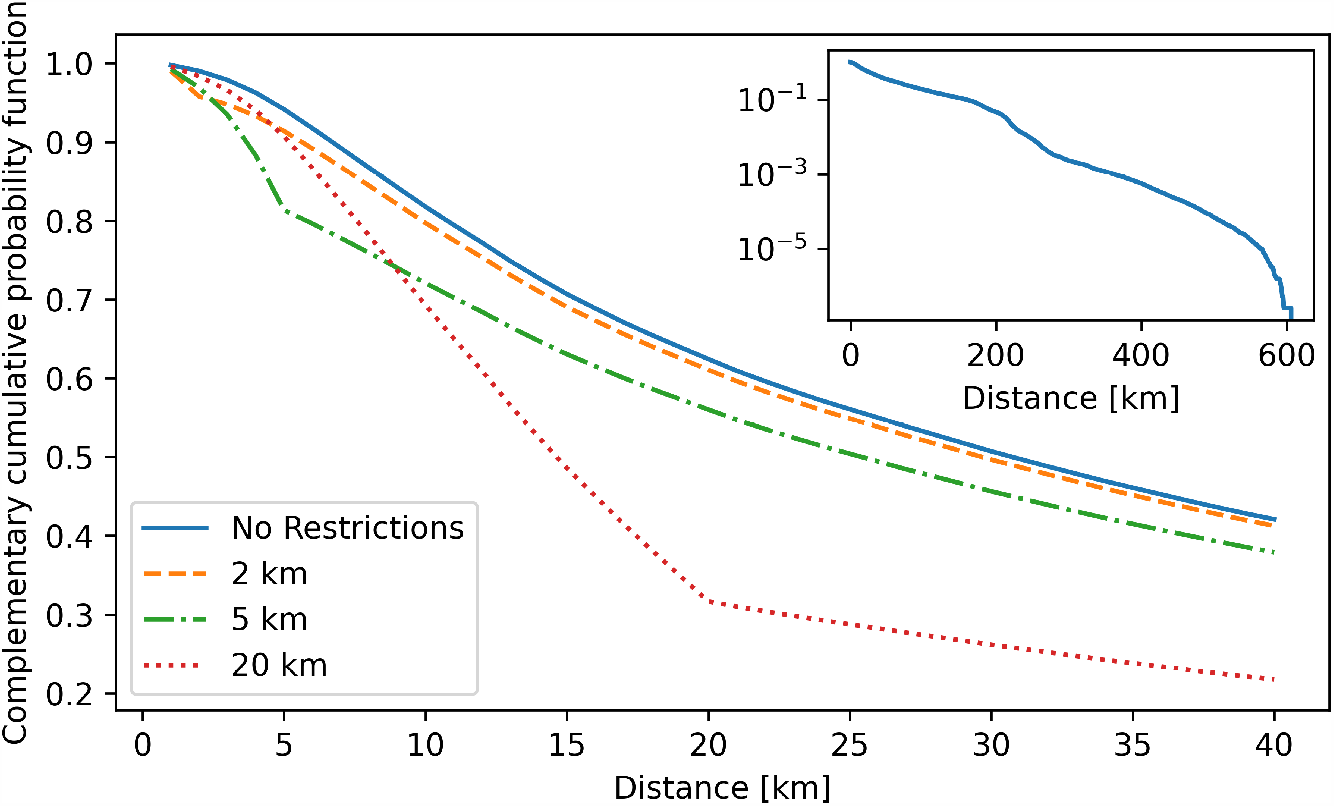
The complementary cumulative proportion of individuals travel distance with the 2km (orange dashed), 5km (green dash-dotted), and 20km (red dotted) restrictions in place for a compliance level of 80%. The original, pre-lockdown data, which are used during periods with no travel restrictions, is shown as blue solid curve. The inset shows the the full range in a semi-log scale.

In addition, we show in Fig. 17 how the travels out of a particular area (here an electoral division in Dublin; red area marked by the arrow) are affected by the different travel restrictions. As the travel radius is reduced, the movement is confined to a more local vicinity. Note that travel beyond the travel radius is still present due to non-compliance.

**Figure 17:**
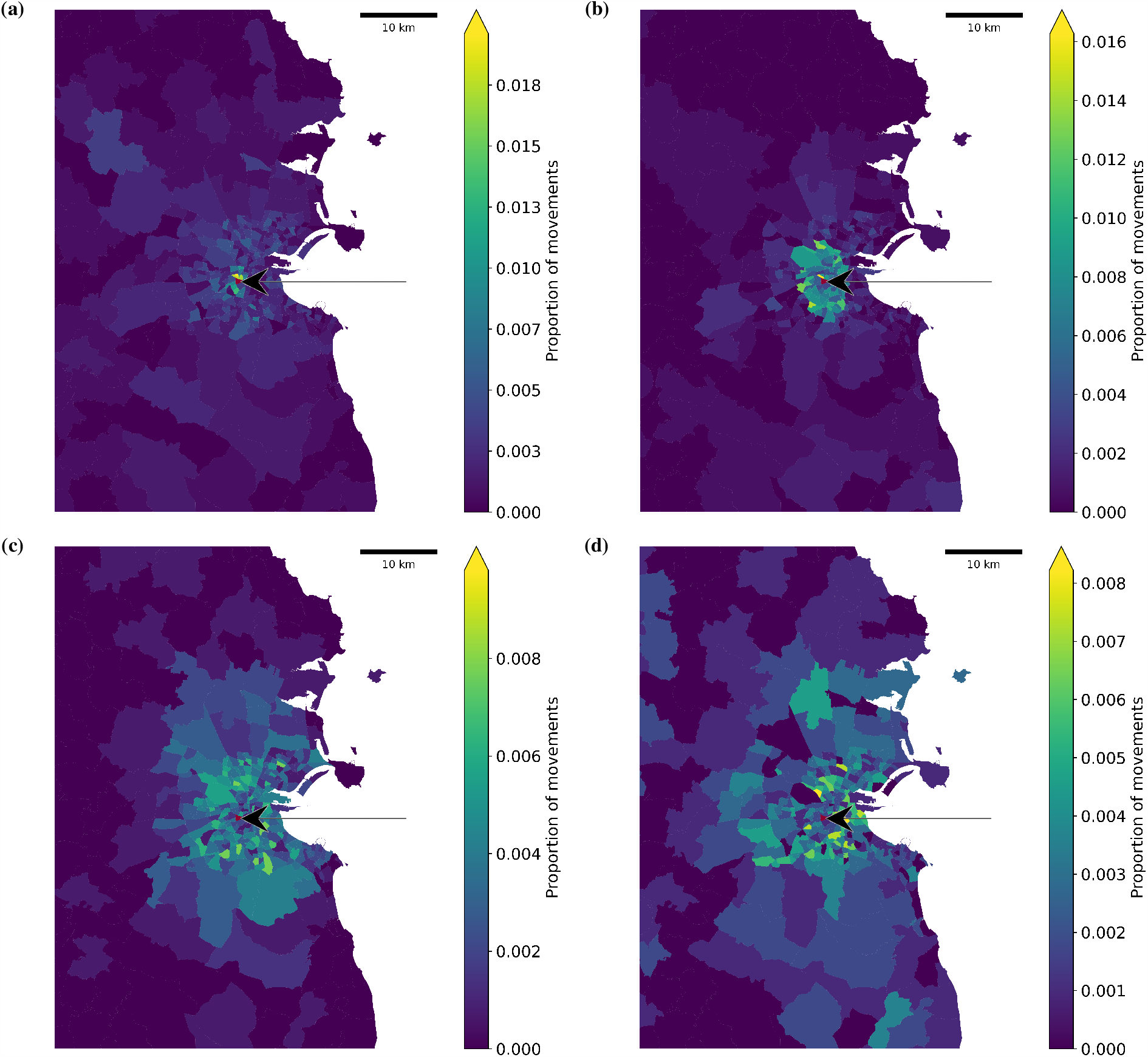
A visual representation of the journeys out of a particular electoral division in the Dublin area (red area marked by the arrow) to surrounding electoral divisions with different restrictions in place with a compliance of 80%. Labels (a), (b), (c), and (d) correspond to travel restrictions of 2km, 5km, 20km, and none, respectively.

## References

[1] Al-Khani, A.M., Khalifa, M.A., AlMazrou, A., Saquib, N., 2020. The SARS-CoV-2 pandemic course in Saudi Arabia: A dynamic epidemiological model. Infectious Disease Modelling 5, 766–771.

[2] Alwan, N.A., Burgess, R.A., Ashworth, S., Beale, R., Bhadelia, N., Bogaert, D., Dowd, J., Eckerle, I., Goldman, L.R., Greenhalgh, T., et al., 2020. Scientific consensus on the COVID-19 pandemic: we need to act now. The Lancet 396, e71–e72.

[3] Amann, A., 2020. How to choose the best strategy for fighting an infectious disease. private communication.

[4] Ames, A.D., Molnar, T.G., Singletary, A.W., Orosz, G., 2020. Safety-critical control of active interventions for COVID-19 mitigation. medRxiv URL: https://www.medrxiv.org/content/early/2020/06/19/2020.06.17.20133264, doi:10.1101/2020.06.17.20133264, arXiv:https://www.medrxiv.org/content/early/2020/06/19/2020.06.17.20133264.full.pdf.

[5] Belik, V., Geisel, T., Brockmann, D., 2011. Natural human mobility patterns and spatial spread of infectious diseases. Phys. Rev. X 1, 011001. doi:10.1103/physrevx.1.011001.

[6] Beverton, R.J., Holt, S.J., 1993. On the dynamics of exploited fish populations. volume 11. Springer Science & Business Media.

[7] Bloomberg, 2021. Covid-19 Vaccine Tracker. https://www.bloomberg.com/graphics/covid-vaccine-tracker-global-distribution/. (Accessed on 01/16/2021).

[8] Blyuss, K.B., Kyrychko, Y.N., 2020. Effects of latency and age structure on the dynamics and containment of COVID-19. medRxiv.

[9] Brockmann, D., Hufnagel, L., 2007. Front propagation in reaction-superdiffusion dynamics: Taming Lévy flights with fluctuations. Phys. Rev. Lett. 98, 178301.

[10] Brockmann, D., Hufnagel, L., Geisel, T., 2006. The scaling laws of human travel. Nature 439, 462–465.

[11] Census 2016 Open Data Site, 2016. Electoral Divisions - CSO Generalised 20m. http://census2016.geohive.ie/datasets/774b6c87f0ec49c48e0e7999fc0e752b_1. (Accessed on 06/17/2020).

[12] Central Statisitcs Office, 2016. Census 2016 Profile 6, Commuting in Ireland. https://www.cso.ie/en/csolatestnews/presspages/2017/census2016profile6-commutinginireland/. (Accessed on 06/17/2020).

[13] Chen, N., Zhou, M., Dong, X., Qu, J., Gong, F., Han, Y., Qiu, Y., Wang, J., Liu, Y., Wei, Y., et al., 2020. Epidemiological and clinical characteristics of 99 cases of 2019 novel coronavirus pneumonia in Wuhan, China: a descriptive study. The Lancet 395, 507–513.

[14] Chowdhury, R., Heng, K., Shawon, M.S.R., Goh, G., Okonofua, D., Ochoa-Rosales, C., Gonzalez-Jaramillo, V., Bhuiya, A., Reidpath, D., Prathapan, S., et al., 2020. Dynamic interventions to control COVID-19 pandemic: a multivariate prediction modelling study comparing 16 worldwide countries. Eur. J. Epidemiol. 35, 389–399.

[15] Christensen, V., Walters, C.J., 2004. Ecopath with Ecosim: methods, capabilities and limitations. Ecol. Modell. 172, 109–139.

[16] Department of the Taoiseach and Department of Health, 2020. Roadmap for reopening society and business, published at: 1 May 2020, last updated: 12 June 2020. https://www.gov.ie/en/news/58bc8b-taoiseach-announces-roadmap-for-reopening-society-and-business-and-u/, https://assets.gov.ie/73722/ffd17d70fbb64b498fd809dde548f411.pdf. (Accessed on 06/17/2020).

[17] Dong, E., Du, H., Gardner, L., 2020. An interactive web-based dashboard to track COVID-19 in real time. Lancet Infect. Dis. 20, 533–534.

[18] Douglas, M., Katikireddi, S.V., Taulbut, M., McKee, M., McCartney, G., 2020. Mitigating the wider health effects of COVID-19 pandemic response. BMJ 369, m1557.

[19] Eurostat, 2018. Nomenclature of Territorial Units for Statistics – NUTS. https://ec.europa.eu/eurostat/web/nuts/background. (Accessed on 06/17/2020).

[20] Grauwin, S., Szell, M., Sobolevsky, S., Hövel, P., Simini, F., Vanhoof, M., Smoreda, Z., Barabási, A.L., Ratti, C., 2017. Identifying and modeling the structural discontinuities of human interactions. Sci. Rep. 7, 46677. doi:10.1038/srep46677.

[21] Health Protection Surveillance Centre (HPSC), 2020a. COVID-19 Cases in Ireland. https://www.hpsc.ie/a-z/respiratory/coronavirus/novelcoronavirus/casesinireland/. (Accessed on 06/22/2020).

[22] Health Protection Surveillance Centre (HPSC), 2020b. Ireland’s COVID-19 Data Hub. https://covid19ireland-geohive.hub.arcgis.com/. (Accessed on 12/22/2020).

[23] HRB project, 2020. COVID19 rapid response programme (grant number: COV19-2020-117). unpublished data of four waves of telephone surveys. (manuscript under review).

[24] Humphries, R., 2020. EpiGraph. https://github.com/rory-humphries/EpiGraph-cpp.

[25] Johns Hopkins University, 2020. COVID-19 dashboard by the Center for Systems Science and Engineering (CSSE) at Johns Hopkins University. https://coronavirus.jhu.edu/map.html. (Accessed on 11/30/2020).

[26] Kermack, W.O., McKendrick, A.G., 1927. A contribution to the mathematical theory of epidemics. Proc. R. Soc. A 115, 700–721.

[27] Killeen, G., Kiware, S., 2020. Why lockdown? simplified arithmetic tools for decision-makers, health professionals, journalists and the general public to explore containment options for the novel coronavirus. medRxiv URL: https://www.medrxiv.org/content/early/2020/04/20/2020.04.15.20066845, doi:10.1101/2020.04.15.20066845, arXiv:https://www.medrxiv.org/content/early/2020/04/20/2020.04.15.20066845.full.pdf.

[28] Kissler, S.M., Tedijanto, C., Goldstein, E., Grad, Y.H., Lipsitch, M., 2020. Projecting the transmission dynamics of ARS-CoV-2 through the postpandemic period. Science 368, 860–868.

[29] Kyrychko, Y.N., Blyuss, K.B., Brovchenko, I., 2020. Mathematical modelling of the dynamics and containment of COVID-19 in Ukraine. Sci. Rep. 10, 19662.

[30] Liu, Z., Magal, P., Seydi, O., Webb, G., 2020. A covid-19 epidemic model with latency period. Infectious Disease Modelling 5, 323–337.

[31] Maier, B.F., Brockmann, D., 2020. Effective containment explains subexponential growth in recent confirmed COVID-19 cases in china. Science 368, 742–746. doi:10.1126/science.abb4557.

[32] Northern Ireland Statistics and Research Agency, 2016. Local Government District: Council area statistics. https://www.nisra.gov.uk/statistics/regional-analysis-and-trends/local-government-district. (Accessed on 06/17/2020).

[33] Pinotti, F., Di Domenico, L., Ortega, E., Mancastroppa, M., Pullano, G., Valdano, E., Boëlle, P.Y., Poletto, C., Colizza, V., 2020. Tracing and analysis of 288 early sars-cov-2 infections outside china: A modeling study. PLOS Med. 17, e1003193.

[34] Prem, K., Liu, Y., Russell, T.W., Kucharski, A.J., Eggo, R.M., Davies, N., Flasche, S., Clifford, S., Pearson, C.A., Munday, J.D., et al., 2020. The effect of control strategies to reduce social mixing on outcomes of the COVID-19 epidemic in Wuhan, China: a modelling study. The Lancet Public Health 5, e261–e270.

[35] Pullano, G., Valdano, E., Scarpa, N., Rubrichi, S., Colizza, V., 2020. Evaluating the effect of demographic factors, socioeconomic factors, and risk aversion on mobility during the COVID-19 epidemic in France under lockdown: a population-based study. The Lancet Digital Health 2, e638–e649.

[36] Schlosser, F., Maier, B.F., Jack, O., Hinrichs, D., Zachariae, A., Brockmann, D., 2020. Covid-19 lockdown induces disease-mitigating structural changes in mobility networks. Proc. Natl. Acad. Sci. U.S.A. 117, 32883–32890. URL: https://www.pnas.org/content/117/52/32883, doi:10.1073/pnas.2012326117, arXiv:https://www.pnas.org/content/117/52/32883.full.pdf.

[37] Simini, F., González, M.C., Maritan, A., Barabási, A.L., 2012. A universal model for mobility and migration patterns. Nature 484, 96–100.doi:10.1038/nature10856.

[38] Song, C., Koren, T., Wang, P., Barabási, A.L., 2010a. Modelling the scaling properties of human mobility. Nat. Phys. 6, 818–823.

[39] Song, C., Qu, Z., Blumm, N., Barabási, A.L., 2010b. Limits of Predictability in Human Mobility. Science 327, 1018–1021. doi:10.1126/science.1177170, arXiv:http://www.sciencemag.org/cgi/reprint/327/5968/1018.pdf.

[40] van den Driessche, P., Watmough, J., 2002. Reproduction numbers and sub-threshold endemic equilibria for compartmental models of disease transmission. Mathematical Biosciences 180, 29 – 48. URL: http://www.sciencedirect.com/science/article/pii/S0025556402001086, doi:https://doi.org/10.1016/S0025-5564(02)00108-6.

[41] World Health Organization, 2020a. Novel coronavirus (2019-ncov). Situation Report 10.

[42] World Health Organization, 2020b. Novel coronavirus (2019-ncov). https://www.who.int/groups/covid-19-ihr-emergency-committee. (Accessed on 11/27/2020).

[43] World Health Organization, 2020c. Novel coronavirus (2019-ncov). Situation Report 51.

[44] Worldometer, 2020. Coronavirus: Ireland. https://www.worldometers.info/coronavirus/country/ireland/. (Accessed on 06/22/2020).

[45] Worldometer, 2020. Coronavirus update (live): 61,177,715 cases and 1,434,894 deaths from COVID-19 virus pandemic - worldometer. https://www.worldometers.info/coronavirus/. (Accessed on 11/27/2020).

